# Cost-Effectiveness of the IMPALA Monitoring System for Hospitalised Children in Low-Resource Settings: A Pragmatic Before-and-After Study

**DOI:** 10.1101/2025.10.31.25339214

**Authors:** Ângela Jornada Ben, Daniel Mwale, Pam Jansen, Owen Mtambo, Niek Versteegde, Eveline Geubbels, Job Calis, Jessica Chikwana, IMPALA study team, Jobiba Chinkhumba, Wendy Janssens

**Affiliations:** Amsterdam Institute of Global Health and Development, Amsterdam, the Netherlands; Department of Health Sciences, Faculty of Science, Vrije Universiteit Amsterdam, Amsterdam, the Netherlands; School of Business and Economics, Vrije Universiteit Amsterdam, Amsterdam, the Netherlands; School of Global and Public Health, Kamuzu University of Health Sciences, Blantyre, Malawi; Training and Research Unit of Excellence, Blantyre, Malawi; GOAL 3, ‘s Hertogenbosch, the Netherlands; Department of Pediatric Intensive Care, Emma Children’s Hospital, Amsterdam University Medical Center, Amsterdam, Netherlands; Department of Pediatrics, College of Medicine, Queen Elizabeth Central Hospital, Blantyre, Malawi

## Abstract

**Background:** Staff shortages, limited training, and inadequate hospital equipment often delay responses to patient deterioration in low-resource settings. The IMPALA continuous monitoring system was developed to support proactive care for critically ill children in such settings. This study evaluated IMPALA cost-effectiveness compared with current practice manual intermittent monitoring from provider and societal perspectives.

**Methods:** We conducted an economic evaluation based on a before-and-after cohort of children (0-180 months) admitted to Zomba Central Hospital (ZCH) and St. Luke’s Hospital (SLH), Malawi (2022-2024), where IMPALA was implemented in high-dependency units (HDUs). Targeted maximum likelihood estimation assessed percentage point (pp) differences in mortality, critical illness events (CIEs), disability-adjusted life years (DALYs), and costs (medical, non-medical, indirect). Incremental cost-effectiveness ratios (ICERs) and cost-effectiveness probabilities were calculated for different willingness-to-pay thresholds.

**Results:** At ZCH paediatric ward, 1,840 pre- and 6,255 post-IMPALA children were included; 248 and 736 were admitted to the HDU. Ward mortality decreased (3.7%–2.8%), with an adjusted 1.9pp reduction (95%CI: - 3.8;-0.6). At ZCH-HDU, mortality slightly increased (8.1%–9.0%), but IMPALA was associated with an adjusted 9.8pp reduction (95%CI: −26.5;5.0), a 47.1pp decrease in CIEs (95%CI: −52.9;-41.8), and 5.4 DALYs averted (95%CI: −14.2;3.1). At SLH paediatric ward, mortality decreased (4.0%–2.1%), with an adjusted 1.6pp reduction (95%CI: −3.2;-0.2), a 25.5pp decrease in CIEs (95% CI: −30.1;-20.9), and 1.0 DALYs averted (95% CI: −1.9;-0.1). Provider and societal costs decreased in both wards, but not in the HDU. IMPALA was dominant in wards and slightly more costly but more effective in the HDU (ICERs -$22.5–$0.4 per life saved). Cost-effectiveness probabilities ranged from 0.8–1.0 in wards and 0.3–1.0 in the HDU.

**Conclusion:** IMPALA was highly cost-effective, reducing mortality by >40%, morbidity by >50%, increasing DALYs averted, shortening hospital stays, and lowering costs, with spillover benefits from HDUs to wards.

**Key messages:** *What is already known on this topic:*

- Continuous monitoring reduces mortality in high-resource settings, but its (cost-)effectiveness in low-resource contexts is uncertain due to staff shortages, unstable power, and limited supplies.

*What this study adds:*

- Tailored for low-resource settings, IMPALA monitoring system saves lives, reduces illness events, and lowers inpatient and societal costs.

*How this study might affect research, practice or policy:*

- Findings support broader implementation of a tailored continuous monitoring system to improve paediatric outcomes in low-resource settings.

## Introduction

Globally, child mortality has declined by 59% in the past three decades.^1^ Despite this progress, child mortality rates remain high in sub-Saharan Africa (SSA), at 75.8 deaths per 1,000 live births in 2019[1], being 19 times higher compared to high-income countries (HIC), and accounting for 55% of global mortality under 5 years of age.[1] Despite increasing access to healthcare, healthcare facilities in low- and middle-income countries (LMICs) generally lack staff and resources to deliver high-quality care. Up to 15% of overall deaths in LMICs can be attributable to poor quality of care and may be prevented by improving care.[2]

An important cause of in-hospital mortality is related to the late detection of deteriorating vital signs.[2,3] This is partly due to reliance on intermittent, manual monitoring, which limits healthcare workers’ ability to promptly respond when a child’s clinical condition worsens.[4–6] Although patient monitoring systems show benefits in high-income settings,[7] the value and feasibility of their implementation in low-resource contexts is not proven. This is important as concerns exist about the performance of these technologies as their beneficial impact may be hindered by high patient-to-staff ratios, limited trained personnel, unreliable electricity, and scarce consumables and parts.[8,9]

To address these challenges, a Malawian and European multidisciplinary team co-designed the Innovative Monitoring in PaediAtrics in Low-resource settings: an Aid to save lives (IMPALA) system – hereafter referred to as IMPALA.[10] The IMPALA consortium includes seven institutes, three of which are based in Africa and one social enterprise (GOAL 3).[10] IMPALA is a multicomponent innovation built around a human-centric service model.[11] It includes a battery-backed automated digital continuous monitoring devices and local server, a tablet-based clinical decision support application (app), staff training and ongoing technical support. The app presents patient data using avatars and a traffic light system to support early detection of clinical deterioration and prioritization of care. Tailored for low-resource settings, the system is robust against heat, moisture, insects, and power outages lasting up to 10 hours. It uses reusable sensors to ensure consistent availability and affordability. The tailored implementation strategy includes technical briefings, emergency and critical care training, guidelines on new care processes, all complemented by an online platform with instructional videos. A ward-level “champions” program reinforces system adoption by assigning staff members responsible for supporting training, promoting knowledge transfer, and resolving technical issues, thereby enhancing ownership, sustainability, and quality improvement.[11]

Mixed-methods assessments have shown that the IMPALA system is well-accepted by both healthcare workers and caregivers.[12–14] Early qualitative evidence from Malawi indicates that the IMPALA system is addressing real barriers,[15] but its (cost)-effectiveness was not yet known. This study assesses the impact of the IMPALA system by evaluating its cost-effectiveness compared to standard care, with mortality and occurrence of critical illness events (CIEs) as primary outcomes and disability-adjusted life years (DALYs) as a secondary outcome, using study setting and real-world data.

## Methods

### Setting and study population

Malawi has an under-five mortality rate of 40 deaths per 1,000 live births.[16] It has a universal healthcare system that provides free access to public care. Zomba Central Hospital (ZCH), one of the country’s four central referral hospitals, serves 830,000 people with approximately 5,000 paediatric admissions annually.[17] The ZCH paediatric ward has 57 beds, including 16 HDU beds, with multiple critically ill children sharing a bed when needed.[15] St. Luke’s Hospital (SLH), is a faith based hospital that serves 90,000 people in the Zomba region and has around 1,000 paediatric admissions annually.^20^ Its paediatric ward has 29 beds including 7 HDU beds. Most paediatric admissions occur in the period January-June, when malaria, anaemia and respiratory infections peak. Both hospitals receive government subsidies, and donor support. SLH supplements its funding through patient contributions. The study population included children aged 0 to 180 months admitted to the ZCH paediatric ward, ZHC-HDU, and to the SLH paediatric ward.

### Study design

This is an economic evaluation based on a retrospective before-and-after cohort study, planned and reported according to the Consolidated Health Economic Evaluation Reporting Standards 2022 (CHEERS) statement.[18] Ethical approval was granted by the University of Malawi College of Medicine Research and Ethics Committee (P.01/22/3552, P.02/22/3575 and P.02/24-0607).

### Data collection

At ZCH, clinical and cost data were collected retrospectively from January 1 to June 30, 2022 (pre-IMPALA cohort), and in two periods after the IMPALA implementation: from January 1 to June 30, 2023, and from January 17 to May 31, 2024. At SLH, data were collected retrospectively for both the pre-IMPALA cohort (February 1, 2022 to January 31, 2023), and the post-IMPALA cohort (February 1, 2023, to January 31, 2024). Clinical data were extracted from paper-based and digital medical charts, and ward registers. Health facility cost data were collected through review of expenditure records and interviews with the hospital manager and financial accountant in 2023 (ZHC) and 2024 (SLH), household costs were obtained through surveys with 150 caregivers during their children’s stay in the ZHC-HDU between January 29 and May 31, 2023. In both hospitals, data were entered via tablet by trained research assistants and stored in the Research Electronic Data Capture (REDCap) system. Although IMPALA was implemented in the HDU, data were collected from both the HDU and paediatric ward to capture spillover effects, as the system was expected to influence care processes and outcomes across the entire ward (**Figure 1**).

**Figure 1.**
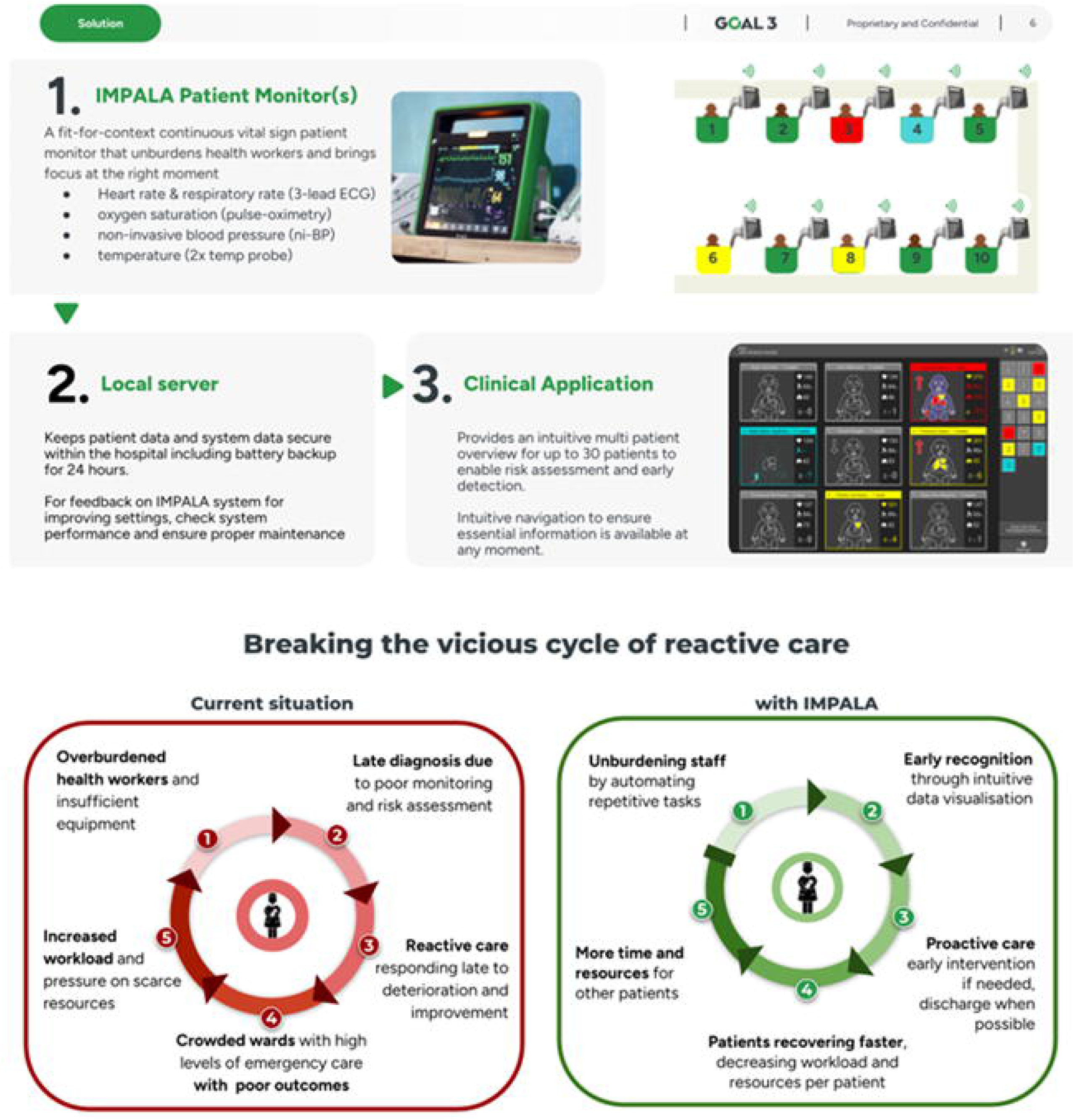
The IMPALA strategy to break the vicious cycle of reactive care. IMPALA aims to shift from reactive to proactive care through five key mechanisms: (1) Reducing staff burden by automating repetitive tasks, allowing health workers to focus on clinical decision-making. (2) Enabling early recognition of abnormal vital signs through real-time monitoring and alerts. (3) Promoting proactive care by streamlining workflows and providing targeted staff training. (4) Preventing critical illness events, thereby reducing the intensity of care and resources required per patient. (5) Optimising time and resource availability, enabling more efficient care for other patients.

Variables collected at admission included age, sex, weight, HIV status, admission date, and diagnosis in both hospitals. Diagnoses were categorized as: respiratory, gastrointestinal, neurological, renal/cardiovascular, systemic or severe infection/inflammation, malaria, malnutrition, hematologic/oncologic, and other (**Supplemental Table 1**). Vital signs (oxygen saturation (SpO2), respiratory rate (RR), heart rate (HR), and temperature) and the Blantyre coma scale (BCS)^21^ were collected only for children admitted to the ZCH-HDU (**Supplemental material**). Vital signs were categorized (abnormal/ normal) based on the World Health Organization guidelines.[19] The BCS scoring ranges from 0 to 5 with scores under 5 considered abnormal.[20]

### Sample size

A sample of 1,353 per cohort was needed to detect a 3% difference in mortality and 197 children per cohort were required for a 10% difference in CIEs (α=0.05, 80% power) based on a pilot including 485 children (pre-IMPALA, n=182, and post-IMPALA, n=303) (**Supplemental material**).[21]

### Standard care

Standard care included intermittent manual monitoring of children’s vital signs by nurses four times a day or more often, depending on the child’s condition.[15] Nurses measured SpO2, RR, HR, temperature, and non-invasive blood pressure (NIBP) if needed.[15] In addition, caregivers informed nurses when observing deterioration.[15]

## IMPALA

IMPALA is a multicomponent clinical monitoring system designed to support early detection of deterioration in hospitalised children. It includes bedside monitoring units that continuously measure vital signs (SpOD, respiratory rate, heart rate, temperature, and non-invasive blood pressure using reusable sensors), a local server for data processing and storage, and a tablet-based app that displays patient data of up to 30 patients simultaneously. The monitors and tablet app generate visual and audible alerts when predefined thresholds are reached, thus allowing rapid response and patient prioritization. The intervention also comprises staff training, implementation support, and ongoing technical assistance (**Figure 1**). Training was delivered in person and supported by ward-level champions who facilitated uptake, sustained use and troubleshooting. Nine and six IMPALA monitoring units were deployed at the HDUs of ZCH and SLH, respectively. Allocation of monitoring was based on clinical assessment by nurses and clinicians. More detailed information is provided in https://www.goal3.org/product.

### Effect outcomes

Mortality and CIEs were the primary effect outcomes. A CIE was defined as any life-threatening event or life-saving intervention occurring during a hospital stay (**Supplemental Table 2**). DALYs were the secondary outcome as they capture both years of life lost due to premature mortality and years lived with disability resulting from CIEs at both the patient and population levels (detailed explanation in **Supplemental material** and **Supplemental Tables 3 and 4**).

### Cost outcomes

This economic evaluation considered both provider and societal perspectives. The provider perspective included the IMPALA costs and direct medical costs (hereafter referred to as inpatient costs). The societal perspective expanded on this by incorporating household direct non-medical costs (e.g., transportation, food) and indirect costs (i.e., productivity losses due to work absenteeism) related to hospital stays (hereafter referred to as societal costs).

IMPALA costs were fixed at US$2.90 per child, estimated as the average system cost across ZCH and SLH as presented in **Supplemental material**. These costs included purchase, implementation, maintenance/repairs, and daily usage, regardless of whether a child was monitored, as the technology would be available even if a child did not need it. Direct medical costs were calculated by multiplying the length of hospital stay in days by the estimated cost per inpatient day, including personnel, medication, food, and other medical equipment. Costs per inpatient day were derived from the ZCH expenditure records and SLH billing (**Supplemental Table 5**).

Direct non-medical and indirect costs were estimated based on the ZCH caregiver survey (**Supplemental Table 6**). Direct non-medical costs were calculated by multiplying the length of hospital stay in days by the average daily expenses incurred by caregivers for transportation, food, accommodation, and other out-of-pocket costs (e.g., over-the-counter medicines, soap). Indirect costs were calculated by multiplying the length of hospital stay in days by the average daily absenteeism cost per caregiver estimated from the caregiver survey (i.e., the self-reported amount of income lost due to missed work per one-day hospital stay).

Facility costs from ZCH were collected in 2023, US dollars ($) and adjusted for Malawi’s 2024 inflation rate.[22] SLH facility costs and direct non-medical and indirect costs were collected in Malawian Kwacha (MWK) converted to $ using the 2024 World Bank PPP rate.[23] Discount rate was not applied due to the short time horizon of six months. Details on cost estimation and unit prices for the cost-effectiveness analysis are in the **Supplemental Table 7**.

### Statistical analysis

A descriptive analysis of complete and missing data was conducted by study location. Mean imputation was used to handle missing values at hospital admission.^29^ Characteristics of children at admission were reported as n (%) for categorical variables and mean with standard deviation (SD) for continuous variables. Given the non-randomized design, cohort differences at admission were tested using t-tests for continuous variables and chi-squared tests for categorical variables.

Targeted maximum likelihood estimation (TMLE) was used to estimate the Average Treatment Effect (ATE) between the pre- and post-IMPALA cohorts in percentage points (pp).[24,25] The ATE represents the average effect of IMPALA on the entire population of children, regardless of their actual use of IMPALA. It reflects system-wide effects such as improved workflows and earlier recognition of deterioration, making it well-suited for evaluating a multicomponent intervention. TMLE was performed in three steps: (1) fitting g-computation models for mortality, CIEs, and DALYs and costs using relevant covariates to estimate ATEs; (2) fitting a propensity score model to match pre- and post-IMPALA cohorts and calculate inverse probability weights; and (3) updating the ATEs based on propensity scores. Cohort balance was assessed via propensity score density plots. Pre-TMLE comparisons were obtained from linear regression models (**Supplemental Figure 1**).

To extrapolate DALYs averted at the population level (i.e., children admitted during the intervention period), two approaches were used: (i) the TMLE-based approach, in which DALYs averted were estimated directly as a single composite outcome from TMLE-derived patient-level estimates; and (ii) the decomposition approach, which uses TMLE patient-level estimates of mortality and morbidity separately and then combines these components to construct the composite DALYs averted (**Supplemental material**). DALYs averted could not be estimated for the ZCH pediatric ward using the TMLE-based approach because CIEs were not measured in this location. In contrast, the decomposition approach allows extrapolation of CIE impacts from the SLH ward, enabling approximation of DALYs for the ZCH ward.

Incremental cost-effectiveness ratios (ICERs) were calculated by dividing the ATE in costs between pre-and post-IMPALA cohorts by the ATE in effects. Bootstrapping with 1,000 replications estimated the joint uncertainty around the ATE in costs and effects, with bootstrapped cost-effect pairs plotted on cost-effectiveness planes (CE-planes).[26] For an easier CE-planes interpretation, the ATEs for mortality, CIEs, and DALYs were multiplied by −1 as a positive value indicates improvement in outcomes. Probabilities of IMPALA being cost-effective compared to standard care were estimated using willingness-to-pay (WTP) thresholds of $0, $224, and $448 per unit of effect gained, based on, respectively, 0, 0.5 and 1 time Malawi’s GDP per capita in 2024.[27] Data analysis was conducted using R (R Core Team, 2024, version 4.2.1).

### Sensitivity analyses

Sensitivity analysis used Coarsened Exact Matching (CEM) to match pre- and post-IMPALA cohorts based on admission variables used in the TMLE model (**Supplemental material**). CEM reduces selection bias through exact matching but may reduce power if matches are limited.[28] Also, the Average Treatment Effect on the Treated (ATET) was estimated using seemingly unrelated regressions consistent with covariates used in TMLE.^36^ Only children monitored by IMPALA were matched to pre-IMPALA controls when this information was available (ZHC-HDU), as ATET measures the effect among treated individuals rather than the entire population. For effect outcomes, we additionally computed E-values to assess the robustness of the observed associations to potential unmeasured confounding.[29] As the primary analysis was conducted on the risk-difference scale (i.e., pp), E-values were approximated by deriving risk ratios from TMLE-estimated counterfactual risks under the intervention (post-IMPALA) and control (pre-IMPALA). Larger E-values indicate greater robustness to unmeasured confounding; in practice, values close to 1 suggest that relatively weak confounding could explain the findings, whereas larger values imply that progressively stronger confounding would be needed to explain away the observed association.[29]

### Data sharing

Anonymized data of this study can be made available for replication purposes.

### Patient and Public Involvement

The IMPALA consortium had an active stakeholder committee including healthcare professionals, hospital directors, parents, and other relevant stakeholders who provided feedback on the study design, implementation, and results, as well as on research derived from the main study.

## Results

### Study population

At ZCH paediatric ward, 1,840 pre- and 6,255 post-IMPALA children (0-180 months) were included; 248 and 736 (0-73 months), respectively, were admitted to HDU. In the post-IMPALA cohort, 446 children were monitored by IMPALA devices (all in the HDU). Children in the post-IMPALA cohort were older, had more multimorbidity, and a higher prevalence of malaria compared to the pre-IMPALA cohort. At SLH, 930 pre-and 1,126 post-IMPALA children were included (0-180 months). Children in the post-IMPALA cohort were also older and had a higher prevalence of HIV, but lower multimorbidity (**Table 1**). At SLH, 29.3% of records had missing information on CIEs (**Supplemental Tables 8 and 9**).

**Table 1.** Characteristics of pre- and post-IMPALA cohorts by location.

| Characteristic | Pre-IMPALA | Post-IMPALA | p-value |
| --- | --- | --- | --- |
| **Paediatric ward, Zomba Central Hospital** | **n=1,840** | **n=6,255** |  |
| Age in months, mean (SD) | 33.2 (33.7) | 40.2 (35.2) | 0.001 |
| Sex, female n (%) | 785 (42.7) | 2748 (43.9) | 0.345 |
| HIV reactive or exposure, n (%) | 35 (1.9) | 76 (1.2) | 0.001 |
| Multimorbidity ($\geq 2$), n (%) | 480 (26.1) | 2171 (34.7) | 0.001 |
| Diagnosis, n (%) |  |  |  |
| Respiratory | 788 (67.6) | 1664 (47.2) | 0.001 |
| Gastrointestinal diseases | 229 (31.2) | 431 (15.6) | 0.001 |
| Neurological | 106 (15.7) | 221 (8.4) | 0.001 |
| Renal/cardiovascular | 57 (8.4) | 91 (3.5) | 0.001 |
| Systemic or severe infection/inflammation | 460 (51.5) | 1454 (46.4) | 0.008 |
| Malaria | 387 (45.3) | 3375 (78.0) | 0.001 |
| Malnutrition | 133 (20.0) | 231 (8.8) | 0.001 |
| Hematologic/oncologic | 117 (17.6) | 784 (29.9) | 0.001 |
| Other | 43 (6.4) | 174 (6.5) | 0.956 |
| **High-Dependency Unit, Zomba Central Hospital** | **n=248** | **n=736** |  |
| Age in months, mean (SD) | 19.9 (15.1) | 23.9 (17) | 0.749 |
| Sex, female n (%) | 92 (37.1) | 329 (44.7) | 0.009 |
| Weight in Kg, mean (SD) | 9.5 (2.9) | 9.7 (3.3) | 0.692 |
| Abnormal oxygen saturation, n (%) | 45 (18.1) | 192 (26.1) | 0.688 |
| Abnormal respiratory rate, n (%) | 144 (58.1) | 470 (63.9) | 0.003 |
| Abnormal heart rate, n (%) | 133 (53.6) | 401 (54.5) | 0.473 |
| Abnormal temperature, n (%) | 80 (32.3) | 257 (34.9) | 0.057 |
| HIV reactive or exposure, n (%) | 8 (3.2) | 27 (3.7) | 0.638 |
| Blantyre coma score, mean (SD) | 5 (0.3) | 4.5 (1) | 0.001 |
| Multimorbidity ($\geq 2$), n (%) | 24 (9.7) | 314 (42.7) | 0.001 |
| Diagnosis, n (%) |  |  |  |
| Respiratory | 158 (63.7) | 351 (47.7) | 0.001 |
| Gastrointestinal diseases | 14 (5.6) | 39 (5.3) | 0.702 |
| Neurological | 3 (1.2) | 119 (16.2) | 0.001 |
| Renal/cardiovascular | 3 (1.2) | 26 (3.5) | 0.137 |
| Systemic or severe infection/inflammation | 33 (13.3) | 118 (16) | 0.001 |
| Malaria | 36 (14.5) | 336 (45.7) | 0.001 |
| Malnutrition | 3 (1.2) | 28 (3.8) | 0.013 |
| Hematologic/oncologic | 3 (1.2) | 109 (14.8) | 0.001 |
| Other | 19 (7.7) | 43 (5.8) | 0.660 |
| **Paediatric ward, St. Luke's Hospital** | **n=930** | **n=1,126** |  |
| Age in months, mean (SD) | 41.9 (36.1) | 43.4 (35.7) | 0.001 |
| Sex, female n (%) | 520 (55.9) | 650 (57.7) | 0.430 |
| HIV infected or exposure, n (%) | 15 (1.6) | 20 (1.8) | 0.001 |
| Multimorbidity ($\geq 2$), n (%) | 581 (62.5) | 656 (58.3) | 0.060 |
| Diagnosis, n (%) |  |  |  |
| Respiratory | 265 (28.5) | 265 (23.5) | 0.012 |
| Gastrointestinal diseases | 5 (0.5) | 1 (0.1) | 0.142 |
| Neurological | 7 (0.8) | 9 (0.8) | 1 |
| Systemic or severe infection/inflammation | 432 (46.5) | 414 (36.8) | 0.001 |
| Malaria | 633 (68.1) | 795 (70.6) | 0.232 |
| Malnutrition | 15 (1.6) | 11 (1.0) | 0.277 |
| Hematologic/oncologic | 94 (10.1) | 113 (10.0) | 1 |
| Other | 200 (21.5) | 275 (24.4) | 0.131 |

Pre- and post-IMPALA represent the cohorts admitted in the period before and after the IMPALA implementation, respectively. SD = Standard Deviation. P-values for cohort differences at admission are based on t-tests for continuous variables and chi-squared tests for categorical variables

### Effect outcomes

At ZCH paediatric ward, observed mortality decreased from 3.7% pre-IMPALA to 2.8% post-IMPALA, with an adjusted 1.9pp reduction (95% CI: −3.8; −0.6). At ZCH-HDU, observed mortality marginally increased from 8.1% to 9.0%, but after adjustment IMPALA was associated with a 9.8pp reduction in mortality (95% CI: −26.5; 5.0), a 47.1pp decrease in the occurrence of CIEs (95% CI: −52.9; −41.8), and 5.4 DALYs averted per admitted child (95% CI: −14.2; 3.1). At SLH paediatric ward, observed mortality decreased from 4.0 to 2.1%, with an adjusted 1.6pp reduction in mortality (95% CI: −3.2; −0.2); a 25.5pp decrease in the occurrence of CIEs (95% CI: −30.1; −20.9), and 1.0 DALYs averted per admitted child (95% CI: −1.9; −0.1) (**Table 2**).

**Table 2.** Effects and cost outcomes by location.

|  | Pre-IMPALA | Post-IMPALA | Comparison of pre- and post-IMPALA<sup>‡</sup><br>(95% CI) | Average Treatment Effect<br>(TMLE adjusted pre-post difference)<br>(95% CI) |
| --- | --- | --- | --- | --- |
| **Paediatric ward, Zomba Central Hospital** |  |  |  |  |
| Effect outcomes | n=1,840 | n=6,255 |  |  |
| Mortality, n (%) | 69 (3.8) | 175 (2.8) | -1.0 (-1.9; 0.04) | -1.9 (-3.8; -0.6) |
| Cost outcomes (\$), mean (SD) | | | | |
| Length of stay | 4.0 (15.1) | 3.5 (12.9) | -0.5 (-1.2; 0.2) | -0.5 (-1.7; 0.7) |
| IMPALA costs | 0 (0) | 2.9 (0) | 2.9 (NA) | 2.9 (NA) |
| Direct medical costs | 129.9 (486.7) | 114.9 (415) | -15.0 (-9.6; 39.4) | -16.3 (-61.2; 21.1) |
| Direct non-medical costs | 257.4 (965.1) | 227.8 (822.9) | -29.6 (-19; 78.2) | -31.7 (-124.9; 42.0) |
| Indirect costs | 3.1 (11.6) | 2.7 (9.9) | -0.4 (-0.2; 0.9) | -0.4 (-1.4; 0.4) |
| Inpatient costs | 129.9 (486.7) | 117.8 (415) | -12.1 (-36.6; 12.4) | -13.1 (-56.9; 22.7) |
| Societal costs | 390.4 (1463.5) | 348.3 (1247.8) | -42.1 (-31.7; 115.8) | -45.1 (-176.9; 62.5) |
| **High-Dependency Unit, Zomba Central Hospital** |  |  |  |  |
| Effect outcome | n=248 | n=736 |  |  |
| Mortality, n (%) | 20 (8.1) | 66 (9.0) | 0.9 (-3.2; 5.0) | -9.8 (-26.5; 5.0) |
| CIEs, n (%) | 222 (89.5) | 326 (44.3) | -45.2 (-51.8; -38.6) | -47.1 (-52.9; -41.8) |
| DALYs, mean (SD) | 5.0 (16.8) | 5.4 (17.4) | 0.4 (-2; 2.9) | -5.4 (-14.2; 3.1) |
| Cost outcomes (\$), mean (SD) | | | | |
| Length of stay | 3.2 (1.9) | 3.0 (3.0) | -0.2 (-0.6; 0.2) | 0.01 (-0.49; 0.51) |
| IMPALA costs | 0 (0) | 2.9 (0) | 2.9 (NA) | 2.9 (NA) |
| Direct medical costs | 102.3 (62.5) | 96.9 (97.2) | -5.4 (-18.4; 7.5) | 0.3 (-20.7; 16.6) |
| Direct non-medical costs | 202.9 (123.9) | 192.1 (192.8) | -10.8 (-36.4; 14.9) | 0.6 (-41; 33) |
| Indirect costs | 2.4 (1.5) | 2.3 (2.3) | -0.1 (-0.4; 0.2) | 0.01 (-0.5; 0.4) |
| Inpatient costs | 102.3 (62.5) | 99.7 (97.2) | -2.6 (-15.5; 10.4) | 3.2 (-19.4; 18.6) |
| Societal costs | 307.6 (187.9) | 294.1 (292.3) | -13.5 (-52.4; 25.4) | 3.8 (-61.7; 52.7) |
| **Paediatric ward, St. Luke's Hospital** |  |  |  |  |
| Effect outcome | n=930 | n=1,126 |  |  |
| Mortality, n (%) | 37 (4.0) | 24 (2.1) | -1.9 (-3.5; -0.2) | -1.6 (-3.2; -0.2) |
| CIEs, n (%) | 535 (78.6), n=681 | 411 (53.2), n=772 | -25.3 (-30.1; -20.5) | -25.5 (-30.1; -20.9) |
| DALYs, mean (SD) | 1.7 (10.0), n=681 | 0.6 (6.2), n=772 | -1.1 (-0.2; -1.9) | -1.0 (-1.9; -0.1) |
| Cost outcomes (\$), mean (SD) | | | | |
| Length of stay | 3.0 (2.9) | 2.9 (2.7) | -0.1 (-0.4; 0.2) | -0.1 (-0.4; 0.1) |
| IMPALA costs | 0 (0) | 2.9 (0) | 2.9 (NA) | 2.9 (NA) |
| Direct medical costs | 95.0 (93.3) | 92.2 (89.6) | -2.8 (-5.2; 10.7) | -3.2 (-11.1; 4.9) |
| Direct non-medical costs | 189.3 (185.9) | 183.8 (178.4) | -5.5 (-10.4; 21.3) | -6.3 (-21.7; 9.9) |
| Indirect costs | 2.3 (2.2) | 2.2 (2.2) | -0.1 (-0.1; 0.3) | -0.1 (-0.3; 0.1) |
| Inpatient costs | 95.0 (93.3) | 95.1 (89.6) | 0.1 (-7.9; 8.1) | -1.9 (-11.4; 6.4) |
| Societal costs | 286.5 (281.5) | 281.1 (270.2) | -5.4 (-18.6; 29.4) | -11.6 (-39.6; 13.7) |

<sup>‡</sup>Linear regression without covariate adjustment. NA = not applicable, as the fixed cost per child did not vary in the data. CIEs = critical illness events during hospital stay (yes/no). DALY = disability-adjusted life-year. ATE = average treatment effect. At Zomba Central Hospital (ZCH) paediatric ward, ATEs were adjusted for age, sex, HIV status, multimorbidity, malaria, and the interaction between cohort and malaria, due to higher malaria frequency in the post-IMPALA period. For the high-dependency unit (HDU), ATEs were adjusted for age, sex, abnormal vital signs, Blantyre Coma Scale, multimorbidity, IMPALA monitoring device, malaria, and the cohort-malaria interaction. At St. Luke's Hospital (SLH) paediatric ward, adjustments were made for age, sex, HIV status, and multimorbidity.

At the population level, DALYs averted were 7,365 for the ZCH paediatric ward (decomposition approach only; cohort size n = 6,255), 4,398 and 3,974 for ZCH-HDU (decomposition and TMLE approaches, respectively; n = 736), and 1,120 and 1,126 for the SLH paediatric ward (decomposition and TMLE approaches, respectively; n = 1,126, **Supplemental material**).

E-values based on approximate risk ratios derived from adjusted pp mortality reductions were 2.8 for the ZCH paediatric ward, 4.1 for ZCH-HDU, and 4.4 for the SLH paediatric ward. For CIEs, corresponding E-values were 3.6 for ZCH-HDU and 2.3 for SLH.

### Length of Stay and Cost outcomes

After the IMPALA implementation, the average hospital length of stay decreased in the paediatric wards at ZCH and SLH, leading to reductions in inpatient and societal costs. In contrast, it was slightly higher in ZCH-HDU, leading to increased costs. The primary cost driver was direct non-medical costs (especially travel costs) in both cohorts at all study sites (**Table 2**). Costs per year of IMPALA implementation were $6,221.4 at ZHC and $4,483.6 at SLH (**Supplemental material**).

### Cost-effectiveness results

At ZCH paediatric ward, ICERs were -$6.8 and -$22.5 per life-saved from the provider and societal perspectives, respectively, indicating that IMPALA was more effective and less costly than standard care (**Table 3**). In the ZCH-HDU, ICERs were $0.3 and $0.4, suggesting that IMPALA was more effective but incurred slightly higher costs per life-saved. In the SLH paediatric ward, ICERs were -$1.2 and -$6.0, showing the same dominance of IMPALA over standard care as in ZCH paediatric ward. The ICERs for CIEs and DALYs followed this same dominance pattern in the paediatric wards and the more-effective-but-more-costly pattern in the HDU.

**Table 3.** Cost-effectiveness results by location.

| Effect outcomes | ΔE (95% CI) | ΔC, \$ (95% CI) | ICER | Cost-effectiveness plane | | | | Probability of cost-effectiveness | | |
| --- | --- | --- | --- | --- | --- | --- | --- | --- | --- | --- |
| | | | | NE | SE | SW | NW | WTP \$0 | WTP \$224 | WTP \$448 |
| Paediatric ward, Zomba Central Hospital |  |  |  |  |  |  |  |  |  |  |
| Mortality %, PP* | 1.9 (0.6; 3.8) | -13.1 (-56.9; 22.7) | -6.8 | 29 | 71 | 0 | 0 | 1.0 | 1.0 | 1.0 |
| Mortality %, SP* | 1.9 (0.6; 3.8) | -45.1 (-176.9; 62.5) | -22.5 | 26 | 74 | 0 | 0 | 1.0 | 1.0 | 1.0 |
| High-Dependency Unit, Zomba Central Hospital |  |  |  |  |  |  |  |  |  |  |
| Mortality %, PP* | 9.8 (-5.0; 26.5) | 3.2 (-18.6; 19.4) | 0.3 | 63 | 30 | 6 | 1 | 0.3 | 1.0 | 1.0 |
| Mortality %, SP* | 9.8 (-5.0; 26.5) | 3.8 (-61.7; 52.7) | 0.4 | 54 | 39 | 6 | 1 | 0.3 | 1.0 | 1.0 |
| CIEs %, PP* | 47.1 (41.8; 52.9) | 3.2 (-18.6; 19.4) | 0.1 | 64 | 36 | 0 | 0 | 0.3 | 1.0 | 1.0 |
| CIEs %, SP* | 47.1 (41.8; 52.9) | 3.8 (-61.7; 52.7) | 0.1 | 55 | 45 | 0 | 0 | 0.3 | 1.0 | 1.0 |
| DALYs, PP* | 5.4 (-3.1; 14.3) | 3.2 (-18.6; 19.4) | 1.0 | 63 | 30 | 6 | 1 | 0.3 | 1.0 | 1.0 |
| DALYs, SP* | 5.4 (-3.1; 14.3) | 3.8 (-61.7; 52.7) | 1.0 | 54 | 38 | 7 | 1 | 0.3 | 1.0 | 1.0 |
| Paediatric ward, St. Luke’s Hospital |  |  |  |  |  |  |  |  |  |  |
| Mortality %, PP* | 1.6 (0.2; 3.2) | -1.9 (-11.4; 6.4) | -1.2 | 33 | 65 | 1 | 0 | 0.8 | 1.0 | 1.0 |
| Mortality %, SP* | 1.6 (0.2; 3.2) | -11.6 (-39.6; 13.7) | -6.0 | 19 | 80 | 1 | 0 | 1.0 | 1.0 | 1.0 |
| CIEs %, PP* | 25.5 (20.9; 30.1) | -1.9 (-11.4; 6.4) | -0.1 | 34 | 66 | 0 | 0 | 0.8 | 1.0 | 1.0 |
| CIEs %, SP* | 25.5 (20.9; 30.1) | -11.6 (-39.6; 13.7) | -0.5 | 19 | 81 | 0 | 0 | 1.0 | 1.0 | 1.0 |
| DALYs, PP* | 1.0 (0.1; 1.9) | -1.9 (-11.4; 6.4) | -2.0 | 33 | 65 | 1 | 0 | 0.8 | 0.9 | 0.9 |
| DALYs, SP* | 1.0 (0.1; 1.9) | -11.6 (-39.6; 13.7) | -12.2 | 19 | 80 | 1 | 0 | 1.0 | 0.9 | 0.9 |

CIEs = critical illness events during hospital stay (yes/no). DALYs = disability-adjusted life-years.
\*Effect outcomes were multiplied by -1 to ensure an interpretable cost-effectiveness plane, as lower mortality indicates a better effect, ensuring a more interpretable cost-effectiveness plane.
PP = provider perspective
SP = societal perspective
$\Delta E$ = Average Treatment Effect (ATE) for effect outcomes.
$\Delta C$ = Average Treatment Effect for costs. CI = confidence interval.
ICER = Incremental Cost-Effectiveness Ratio (\$ per unit of effect gained).
NE = Northeast (IMPALA is more costly and more effective than standard care).
SE = Southeast (IMPALA is less costly and more effective than standard care) -> cost-saving

In the paediatric wards of both hospitals, most bootstrapped cost-effect pairs for all outcomes (65-81%) were located in the southeast (SE) quadrant of the CE-plane (**Table 3**), indicating that in these settings, IMPALA was generally more effective and less costly than standard care. The probability of IMPALA being cost-effective compared to standard care ranged from 0.8 to 1.0 for all outcomes across WTP thresholds per unit of effect gained, indicating strong evidence that IMPALA is cost-effective at thresholds corresponding to 0, 0.5, and 1 times Malawi’s GDP per capita.

In the HDU, most cost-effect pairs for all outcomes (54-64%) were in the northeast (NE) quadrant, suggesting that, among children admitted to the HDU, IMPALA was more effective but also more costly than standard care. The probability of IMPALA being cost-effective compared to standard care ranged from 0.3 to 1.0 for all outcomes across WTP thresholds per unit of effect gained; as WTP increases above zero, the greater health gains with IMPALA are valued more highly than the extra costs, leading to high probabilities of cost-effectiveness. **Supplemental Figures 1, 2, and 3** illustrate **Table 3** results, including the CE-planes for all outcomes and the three locations individually. All ICERs (red dots) and most bootstrapped cost-effect pairs (blue dots) of the paediatric wards in the two hospitals fell in the SE quadrants; i.e., simulation scenarios indicated a high degree of certainty around results from both provider and societal perspectives. Results of the sensitivity analyses were in the same direction as those of the main analysis (**Supplemental Table 10**).

Findings were presented and discussed with stakeholders, particularly healthcare providers, whose input contributed to a more accurate description of the IMPALA intervention, its relevance to daily clinical practice, and the practical interpretation of the cost-effectiveness findings in terms of costs and outcomes illustrated in the cost-effectiveness plane, and DALYs averted, with a focus on practical implications at populational level.

### Discussion Main findings

This is the first study to evaluate the cost-effectiveness of a continuous monitoring system tailored to low-resource settings characterized by high patient-to-staff ratios and frequent power outages. IMPALA implementation in paediatric wards was associated with both significantly improved outcomes and cost savings. Mortality rates decreased by 1.6-1.9 pp, a relative reduction of more than 40%. Total costs from both a provider and societal perspective decreased due to shorter hospital stays, and a likely improvement in workflow. In the HDU, the reduction in mortality was not statistically significant, possibly due to limited sample size, but the risk of CIEs fell by more than 50%, corresponding to an estimated 5 DALYs averted per admitted child. HDU costs slightly increased owing to similar lengths of stay pre- and post-IMPALA. Sensitivity analyses yielded consistent results.

When extrapolated to the population level, IMPALA would avert 7,365 DALYs at ZCH among 6,255 children, at an annualized investment of $6,221.4. This represents a conservative lower-bound estimate accounting for potential overlap between paediatric ward and HDU cohorts. The upper-bound estimate would be 11,763 DALYs averted in 6,991 children (7,365 from the paediatric ward plus 4,398 from the HDU; 6,255 plus 736 children, respectively). At SLH, 1,120 DALYs would be averted among 1,126 children, at an annualized investment of $4,483.6.

Cost-effectiveness analysis showed that IMPALA was consistently dominant in paediatric wards, being both more effective and less costly across all outcomes, with ICERs well below Malawi’s WTP thresholds from both health provider and societal perspectives. This may reflect spillover effects from its implementation in the HDU, possibly because the IMPALA system reduces nurses’ time spent on vital signs monitoring, thereby increasing available time to care for other children[12]. Indeed, prior mixed-methods work suggests that IMPALA reduces time spent on monitoring and improves care processes [12]. However, spillover effects were not evaluated in this study, therefore this hypothesis was directly tested. In HDUs, new technologies usually raise costs as a trade-off for reducing mortality, preventing CIEs, and averting DALYs. By contrast, IMPALA improved outcomes in HDUs with only a modest increase in costs.

### Comparison with the literature

Our findings expand on existing evidence from high-resource settings. Based on before-and-after studies and one randomized controlled trial in HICs, a recent meta-analysis found that the use of monitoring systems was associated with a 15% relative reduction in mortality risk compared to standard care (pooled RR 0.85, 95% CI 0.72; 0.99) and 17% relative reduction in clinical deterioration events (pooled RR 0.83, 95% CI 0.62; 1.11).[7] The substantially higher reductions found in our study emphasize the much greater scope for improvement in low-resource settings.

IMPALA is a comprehensive system rather than a monitoring device alone, which limits direct comparisons with prior studies. Only a few economic evaluations exist of similar multicomponent monitoring systems, which differ in study population, cost methodology, modelling approach, and WTP thresholds. Nevertheless, our findings are consistent with available evidence. Guinness et al., using a Delphi consensus among experts, estimated the costs per patient of the Essential Emergency and Critical Care approach (EECC, i.e., vital signs intermittent monitoring, oxygen therapy, and intravenous fluids) in Tanzania and Kenya.[30] The EECC cost per patient day ranged from $1 to $33 in Tanzania, and $2 to $37 in Kenya – generally higher than the IMPALA cost of $3 per patient (**Supplemental material**), regardless of length of stay. Our fixed-rate approach helps avoid additional costs for patients requiring extended care. Similar to our results, Shah et al. show a high probability (0.9) of the EECC being cost-effective for critically ill adult patients compared to standard care relative to a WTP threshold of $101 per DALY averted in Tanzania.[31]

### Limitations

This study has some limitations. First, the non-randomized design is prone to bias due to unobserved confounders. We addressed observed confounders using TMLE, which provides consistent estimates as long as the outcome or propensity score model is correctly specified (but not necessarily both). Propensity score suggested that patients had comparable characteristics between cohorts, with sufficient overlap to support valid estimation of treatment effects. We assessed potential unmeasured confounding using E-values, which indicate the minimum strength of association that an unmeasured confounder would need to have with both exposure and outcome to fully explain the observed associations.[29] The E-values indicated that such confounding would need to be at least moderately strong. Second, the limited number of IMPALA devices meant that some children lacked monitoring. Although models were adjusted for device access, the effect a full roll-out may be larger. Third, inpatient costs were estimated using a simplified bed-day approach, assuming uniform resource use, which may not capture variability in patient needs; patient-level cost data would be more precise. Fourth, DALYs effect may be underestimated, as disability weights from a limited set of chronic diseases were used and the YLD component was restricted to the duration of hospitalisation in HDU or paediatric wards. This means that our DALYs estimates capture YLL plus YLD related to in-hospital health loss only, and do not include post-discharge disability or long-term sequelae. Incorporating long-term sequelae through modelling would allow a more comprehensive assessment of the IMPALA’s impact on disease burden. Fifth, the ability to detect effects on mortality may have been limited by low power in the ZCH-HDU analysis, lower-than-expected mortality in the pre-IMPALA SLH cohort, and restriction of outcomes to the in-hospital period, which may also lead to overestimation of effects if post-discharge deaths were not captured. In contrast, the study was adequately powered to detect differences in CIEs.

### Implications for clinical practice and decision making

This study applied advanced statistical methods to data from three diverse settings. We thereby accounted for real-world data complexities and enhanced the generalizability of our results. Findings indicate that IMPALA is consistently dominant in paediatric wards, reducing mortality, CIEs, and DALYs at low cost, with additional benefits in the HDU where CIEs fell by nearly 50% at only modest cost increases. Implementing IMPALA in HDUs of central and district hospitals under real-world conditions appears cost-saving and life-saving for critically ill children, with positive spillover effects on general paediatric wards.

### Conclusion

This observational study showed that implementing the IMPALA continuous monitoring system, tailored to low-resource settings, together with its implementation strategy, was associated with a highly cost-effective impact: a reduction in overall mortality of more than 40%, a reduction in morbidity of over 50%, more DALYs averted, shorter hospital stays, and lower overall costs.

## Authors’ Contributions

ÂJB was responsible for Methodology, Formal analysis, Investigation, Writing - Original Draft. GR for Writing - Formal analysis, Writing - Review & Editing. DM, OM, PJ, and EG for Investigation, Data curation, Writing - Review & Editing. NV for Investigation, Software, Writing - Review & Editing. JCa for Conceptualization, Software, Project administration, Funding acquisition, Writing - Review & Editing. JCh for Conceptualization, Methodology, Formal analysis, Investigation, Writing - Review & Editing. JeC for Writing - Review & Editing. WJ for Conceptualization, Methodology, Investigation, Resources, Funding acquisition, Writing - Review & Editing, Supervision. All authors read and approved the manuscript.

## Data Availability

Anonymized data of this study can be made available for replication purposes.

## Acknowledgment

We thank Gabriela Riebeek and William Nkhono for excellent research assistance. We thank the IMPALA scientific advisory board (EM Molyneux, MA Ansermino, A Argent, M Hamaluba, M Heys, V Naanyu) for their continuous voluntary support. We further thank the hospital staff, the patients and their guardians for their participation in this study.

