## supplemental material for "Cost-Effectiveness of the IMPALA Monitoring System for Hospitalised Children in Low-Resource Settings: A Pragmatic Before-and-After Study"

1. **List of investigators**

**IMPALA study team**

Collaborators in the IMPALA study team include: Job Calis MD PhD, Amsterdam Institute for Global Health and Development, Amsterdam, The Netherlands; Christopher Pell PhD, Amsterdam Institute for Global Health and Development, Amsterdam, The Netherlands; Wendy Janssens PhD Prof, Amsterdam Institute for Global Health and Development, The Netherlands; Ângela Jornada Ben MD PhD, Amsterdam Institute for Global Health and Development, The Netherlands; Daniella Brals PhD, Amsterdam Institute for Global Health and Development, The Netherlands; Mark Hoogendoorn PhD Prof, Amsterdam Institute for Global Health and Development, The Netherlands; Michaël Boele van Hensbroek MD PhD Prof, Amsterdam Institute for Global Health and Development,The Netherlands; Job van Woensel MD PhD Prof, Emma Childrens’ Hospital of the Amsterdam University Medical Centers, The Netherlands; Natacha Berbers MSc, Amsterdam Institute for Global Health and Development, The Netherlands; Nina Meels MSc, Amsterdam Institute for Global Health and Development, The Netherlands; Valeria Cristofoli MSc, Amsterdam Institute for Global Health and Development, The Netherlands; Keerthana Raghavan MSc, Amsterdam Institute for Global Health and Development, The Netherlands; Niek Versteegde MD, GOAL 3 B.V., ’s-Hertogenbosch, The Netherlands; Bart Bierling MSc, GOAL 3 B.V., ’s-Hertogenbosch, The Netherlands; Eline Pieck-KleinJan MSc, GOAL 3 B.V., ’s-Hertogenbosch, The Netherlands; Eveline Geubbels PhD, GOAL 3 B.V., ’s-Hertogenbosch, The Netherlands; Lieke de Mare MSc, GOAL 3 B.V., ’s-Hertogenbosch, The Netherlands; Lennart Blom MD PhD, GOAL 3 B.V., ’s-Hertogenbosch, The Netherlands; Michael Levin MD PhD Prof, Section of Paediatric Infectious Disease, Imperial College London, London W2 1PG, UK; Aubrey Cunnington MD PhD, Section of Paediatric Infectious Disease, Imperial College London, London W2 1PG, UK; Myrsini Kaforou PhD; Section of Paediatric Infectious Disease, Imperial College London, London W2 1PG, UK; Clare Wilson MD, Section of Paediatric Infectious Disease, Imperial College London, London W2 1PG, UK; Diego Estrada-Rivadeneyra PhD, Section of Paediatric Infectious Disease, Imperial College London, London W2 1PG, UK; Shea Hamilton PhD, Section of Paediatric Infectious Disease, Imperial College London, London W2 1PG, UK; Victoria Wright PhD, Section of Paediatric Infectious Disease, Imperial College London, London W2 1PG, UK; Jonathan Sturgeon PhD, Section of Paediatric Infectious Disease, Imperial College London, Norfolk Place, London W2 1PG, UK; Grieves Mang’anda Jr MPH, Kamuzu University of Health Sciences, Malawi; Marrianne Kasiya MPH, Kamuzu University of Health Sciences, Malawi; Arox W. Kamng’ona PhD Prof, Kamuzu University of Health Sciences, School of Life Sciences and Allied Health Professions, Blantyre, Malawi; Daniel Mwale MSc, Kamuzu University of Health Sciences, Malawi; David Chaima PhD, Kamuzu University of Health Sciences, Malawi; Jenala Njirammadzi - Maleta MD, Kamuzu University of Health Sciences, Malawi; Josephine Langton MBChB Ass Prof, Kamuzu University of Health Sciences, Malawi; Jacquline Msefula MSc, Kamuzu University of Health Sciences, Malawi; James Makina MD, Kamuzu University of Health Sciences, Malawi; Jobiba Chinkhumba PhD, Kamuzu University of Health Sciences, Malawi; Lucinda Manda Taylor PhD; Kamuzu University of Health Sciences, Malawi; William Nkhono MSc, Kamuzu University of Health Sciences, Malawi; Margret Havara BSc, Kamuzu University of Health Sciences, Malawi; Alick Vweza PhD, Malawi University of Business and Applied Sciences, Malawi; Brenald Dzonzi BSc, Malawi University of Business and Applied

Sciences, Malawi; Chimwemwe Msosa PhD, Malawi University of Business and Applied Sciences, Malawi; Christina Chiziwa BSc, Malawi University of Business and Applied Sciences, Malawi; Lezzie Chirambo BSc, Malawi University of Business and Applied Sciences, Malawi; Theresa Mkandawiri PhD Prof, Malawi University of Business and Applied Sciences, Malawi; María Villalobos-Quesada PhD, National eHealth Living Lab, Public Health and Primary Care Department, Leiden University Medical Center, The Netherlands; Margot Rakers MD, National eHealth Living Lab, Public Health and Primary Care Department, Leiden University Medical Center, The Netherlands; Foteini Klapsaki Bach, National eHealth Living Lab, Public Health and Primary Care Department, Leiden University Medical Center, The Netherlands; Kamija Phiri MD PhD Prof, Training Research Unit of Excellence, Malawi; Alice Likumbo BSc, Training Research Unit of Excellence, Malawi; Jessica Chikwana MD, Department of Paediatrics, Zomba Central Hospital, Malawi; Glory Mzembe MD, Training Research Unit of Excellence, Malawi; Mary Magoya MSc, Training Research Unit of Excellence, Malawi; Timothy Rambiki MD, Training Research Unit of Excellence, Malawi; Martin Mwangi PhD, Training Research Unit of Excellence, Malawi; Owen Mtambo PhD, Training Research Unit of Excellence, Malawi; Christopher Nkhata, Training Research Unit of Excellence, Malawi; Patrick Chalira, Training Research Unit of Excellence, Malawi.

2. **Diagnosis definition**

Diagnoses were collected from paper-based medical records via tablet and stored in the Research Electronic Data Capture (REDCap) system. Diagnoses were categorized into 1=Hematologic/Oncologic, 2=Respiratory, 3=Gastrointestinal, 4=Neurological, 5=Renal/Cardiovascular, 6=Other, 7=Systemic or Severe Infection/Inflammation, 8=Malaria, and 9=Malnutrition as shown in **Supplemental Table 1**.

**Supplemental Table 1. Diagnosis definition**

| **dis_diagnosis_1** | **dis_cardiovascular_1** | **Category** | **Number** |
| --- | --- | --- | --- |
| cardiovascular | arrhythmia | Renal-cardiovascular | 5 |
|  | cardiac arrest | Renal-cardiovascular | 5 |
|  | cardiac ischaemia | Renal-cardiovascular | 5 |
|  | congestive heart failure | Renal-cardiovascular | 5 |
|  | central artery vasculitis | Renal-cardiovascular | 5 |
|  | endocarditis | Systemic-severe infection-inflammation | 7 |
|  | myocarditis | Renal-cardiovascular | 5 |
|  | pericarditis | Renal-cardiovascular | 5 |
|  | peripheral artery vasculitis | Renal-cardiovascular | 5 |
|  | acute rheumatic fever | Renal-cardiovascular | 5 |
|  | newly diagnosed congenital heart condition | Renal-cardiovascular | 5 |
|  | hypertension | Renal-cardiovascular | 5 |
|  | other cardiovascular | Renal-cardiovascular | 5 |
| **dis_diagnosis_1** | **dis_git_1** |  |  |
| gastrointestinal | Moderate/severe diarrhoea (<2 weeks) | Gastrointestinal | 3 |
|  | Moderate/severe vomiting | Gastrointestinal | 3 |
|  | necrotising enterocolitis | Gastrointestinal | 3 |
|  | non-specific abdominal pain | Gastrointestinal | 3 |
|  | cholera | Gastrointestinal | 3 |
|  | bloody diarrhoea | Gastrointestinal | 3 |
|  | non-specific abdominal pain (duplicated) | Gastrointestinal | 3 |
|  | proven intestinal parasitic infection | Gastrointestinal | 3 |
|  | chronic diarrhoea (>2 weeks) | Gastrointestinal | 3 |
|  | other GIT | Gastrointestinal | 3 |
| **dis_diagnosis_1** | **dis_haematocardial_1** |  |  |
| hematologic | deep vein thrombosis | Hematologic/Oncologic | 1 |
|  | pulmonary embolism | Hematologic/Oncologic | 1 |
|  | sickle cell crisis | Hematologic/Oncologic | 1 |
|  | severe anemia <5g/dl | Hematologic/Oncologic | 1 |
|  | moderate anemia 5-9 g/dl | Hematologic/Oncologic | 1 |
|  | other haematological | Hematologic/Oncologic | 1 |
| **dis_diagnosis_1** | **dis_inflammatory_1** |  |  |
| inflammatory | ADEM (Acute disseminated encephalomyelitis (ADEM) | Neurological | 4 |
|  | ANCA-assosciated vasculitis | Systemic-severe infection-inflammation | 7 |
|  | crohn’s disease | Systemic-severe infection-inflammation | 7 |
|  | dermatomyositis | Systemic-severe infection-inflammation | 7 |
|  | familial mediterranean fever | Systemic-severe infection-inflammation | 7 |
|  | guillain-barre syndrome | Neurological | 4 |
|  | henoch-schonlein purpura | Systemic-severe infection-inflammation | 7 |
|  | Juvenile idiopathic arthritis (oligoarticular) | Systemic-severe infection-inflammation | 7 |
|  | Juvenile idiopathic arthritis (polyarticular) | Systemic-severe infection-inflammation | 7 |
|  | Juvenile idiopathic arthritis (systemic) | Systemic-severe infection-inflammation | 7 |
|  | kawasaki disease | Systemic-severe infection-inflammation | 7 |
|  | macrophage activation syndrome | Systemic-severe infection-inflammation | 7 |
|  | PFAPA( periodic fever, aphthous stomatitis, pharyngitis, adenitis) | Systemic-severe infection-inflammation | 7 |
|  | reactive arthritis | Systemic-severe infection-inflammation | 7 |
|  | rheumatoid arthritis | Systemic-severe infection-inflammation | 7 |
|  | sarcoidosis | Systemic-severe infection-inflammation | 7 |
|  | stills disease | Systemic-severe infection-inflammation | 7 |
|  | steven johnson syndrome | Systemic-severe infection-inflammation | 7 |
|  | systemic lupus erythematosus | Systemic-severe infection-inflammation | 7 |
|  | systemic scleroderma | Systemic-severe infection-inflammation | 7 |
|  | other periodic fever syndromes | Systemic-severe infection-inflammation | 7 |
|  | undifferentiated inflammatory bowel disease | Systemic-severe infection-inflammation | 7 |
|  | covid related inflammation | Systemic-severe infection-inflammation | 7 |
|  | other autoinflammatory | Systemic-severe infection-inflammation | 7 |
| **dis_diagnosis_1** | **dis_lower_respiratory_tract_1** |  |  |
| lower respiratory tract | asthma exacerbation | Respiratory | 2 |
|  | bronchiolitis | Respiratory | 2 |
|  | empyema | Respiratory | 2 |
|  | exacerbation of COPD | Respiratory | 2 |
|  | pleural effusion | Respiratory | 2 |
|  | pneumonia | Respiratory | 2 |
|  | pneumothorax | Respiratory | 2 |
|  | undefined LRTI | Respiratory | 2 |
|  | viral-induced wheezing | Respiratory | 2 |
|  | chronic cough (>3 weeks) | Respiratory | 2 |
|  | other LRT | Respiratory | 2 |
| **dis_diagnosis_1** | **dis_neurological_1** |  |  |
| neurological | neurological abscess | Neurological | 4 |
|  | encephalitis | Neurological | 4 |
|  | meningitis | Neurological | 4 |
|  | seizure | Neurological | 4 |
|  | stroke/CVA | Neurological | 4 |
|  | coma | Neurological | 4 |
|  | cerebral malaria | Malaria | 8 |
|  | other neurological | Neurological | 4 |
| **dis_diagnosis_1** | **dis_pathogen_syndromes_1** |  |  |
| pathogen syndromes | covid 19 | Systemic-severe infection-inflammation | 7 |
|  | EBV/glandular fever | Systemic-severe infection-inflammation | 7 |
|  | flu-like illness | Systemic-severe infection-inflammation | 7 |
|  | hand-foot-mouth disease | Systemic-severe infection-inflammation | 7 |
|  | hepatitis A | Systemic-severe infection-inflammation | 7 |
|  | HSV stomatitis | Systemic-severe infection-inflammation | 7 |
|  | lyme disease | Systemic-severe infection-inflammation | 7 |
|  | malaria | Malaria | 8 |
|  | measles | Systemic-severe infection-inflammation | 7 |
|  | mumps | Systemic-severe infection-inflammation | 7 |
|  | pertussis | Systemic-severe infection-inflammation | 7 |
|  | roseola/HHV6 | Systemic-severe infection-inflammation | 7 |
|  | tuberculosis | Systemic-severe infection-inflammation | 7 |
|  | scarlet fever | Systemic-severe infection-inflammation | 7 |
|  | staphylococcal scalded skin syndrome | Systemic-severe infection-inflammation | 7 |
|  | varicella (chickenpox) | Systemic-severe infection-inflammation | 7 |
|  | pneumocystis carinii pneumonia | Systemic-severe infection-inflammation | 7 |
|  | scabies | Systemic-severe infection-inflammation | 7 |
|  | HIV | Systemic-severe infection-inflammation | 7 |
|  | typhoid fever | Systemic-severe infection-inflammation | 7 |
|  | **other pathogen syndromes** | **Systemic-severe infection-inflammation or Malaria** | **7 or 8** |
| **dis_diagnosis_1** | **dis_sepsis_syndromes_1** |  |  |
| sepsis syndromes | bacteraemia | Systemic-severe infection-inflammation | 7 |
|  | febrile neutropenia | Systemic-severe infection-inflammation | 7 |
|  | purpura fulminans | Systemic-severe infection-inflammation | 7 |
|  | septic shock | Systemic-severe infection-inflammation | 7 |
|  | sepsis + organ dysfunction | Systemic-severe infection-inflammation | 7 |
|  | toxic shock syndromes | Systemic-severe infection-inflammation | 7 |
|  | other sepsis | Systemic-severe infection-inflammation | 7 |
| **dis_diagnosis_1** | **dis_soft_tissue_1** |  |  |
| soft tissue / skin | abscess | Systemic-severe infection-inflammation | 7 |
|  | cellulitis | Systemic-severe infection-inflammation | 7 |
|  | impetigo | Systemic-severe infection-inflammation | 7 |
|  | infected eczema | Systemic-severe infection-inflammation | 7 |
|  | lymphadenitis | Systemic-severe infection-inflammation | 7 |
|  | necrotising fasciitis | Systemic-severe infection-inflammation | 7 |
|  | periorbital/orbital cellulitis | Systemic-severe infection-inflammation | 7 |
|  | wound infection | Systemic-severe infection-inflammation | 7 |
|  | rash (undifined cause) | Systemic-severe infection-inflammation | 7 |
|  | other soft tissue / skin | Other | 6 |
| **dis_diagnosis_1** | **dis_surgical_1** |  |  |
| surgical | appendicits | Systemic-severe infection-inflammation | 7 |
|  | bowel obstruction | Other | 6 |
|  | mesenteric adenitis | Systemic-severe infection-inflammation | 7 |
|  | pancreatitis | Systemic-severe infection-inflammation | 7 |
|  | peritonitis | Systemic-severe infection-inflammation | 7 |
|  | elective surgery | Other | 6 |
|  | other surgical | Other | 6 |
| **dis_diagnosis_1** | **dis_undifferentiated_fever_1** |  |  |
| undifferentiated fever | febrile convulsion | Systemic-severe infection-inflammation | 7 |
|  | fever without source | Systemic-severe infection-inflammation | 7 |
|  | fever unknown origin | Systemic-severe infection-inflammation | 7 |
|  | post procedural fever | Systemic-severe infection-inflammation | 7 |
|  | other fever | Systemic-severe infection-inflammation | 7 |
| **dis_diagnosis_1** | **dis_urinar_tract_1** |  |  |
| urinary tract | haemolytic uremic syndrome | Systemic-severe infection-inflammation | 7 |
|  | pyelonephritis | Systemic-severe infection-inflammation | 7 |
|  | UTI (urinary tract infection) | Renal-cardiovascular | 5 |
|  | nephrotic/nephritic syndrome | Renal-cardiovascular | 5 |
|  | other urinary | Renal-cardiovascular | 5 |
| **dis_diagnosis_1** | **dis_urti_ent_1** |  |  |
| upper respiratory tract infection | abscess | Respiratory | 2 |
|  | croup | Respiratory | 2 |
|  | mastoiditis | Other | 6 |
|  | otitis media | Other | 6 |
|  | pharyngitis | Respiratory | 2 |
|  | sinusitis | Other | 6 |
|  | stomatitis / lip infection | Other | 6 |
|  | tonsillitis | Respiratory | 2 |
|  | urti (non specific) | Respiratory | 2 |
|  | oral trush | Other | 6 |
|  | other URTI/ENT | Other | 6 |
| **dis_diagnosis_1** | **dis_other_infections_1** |  |  |
| other infections | CLABSI (Central line-associated bloodstream infections) | Systemic-severe infection-inflammation | 7 |
|  | conjunctivitis | Other | 6 |
|  | other infections | Systemic-severe infection-inflammation | 7 |
| **dis_diagnosis_1** | **dis_other_conditions_1** |  |  |
| other conditions | accidents/bites | Other | 6 |
|  | poisoning | Other | 6 |
|  | malnutrition - kwash&marasm | SAM/Malnutrition | 9 |
|  | hypoglycemia | Other | 6 |
|  | jaundice | Other | 6 |
|  | undetermined disease | Other | 6 |
|  | diabetes/DKA | Other | 6 |
|  | malnutrition - kwash | SAM/Malnutrition | 9 |
|  | congenital conditions/syndr | Other | 6 |
|  | no diagnosis | Other | 6 |
|  | malnutrition - marasmus | SAM/Malnutrition | 9 |
|  | other | Other | 6 |

3. **Vital signs**

Vital signs (oxygen saturation (SpO2), respiratory rate (RR), heart rate (HR), and temperature) and the Blantyre Coma Scale^14^ were collected only for children admitted to the Zomba Central Hospital High-Dependency Unit, aged between 28 days and 72 months. Vital signs were categorized as normal or abnormal based on age threshold values determined by the World Health Organization guideline (<https://www.who.int/publications-detail-redirect/978-92-4-154837-3>). Threshold values are as follows: 1) SpO2: Any measurement below 92% was considered life-threatening or abnormal; 2) RR: for infants ≤ 2 months old, normal rates were between 40 and 60 breaths per minute; for infants between 2 and 11 months old, between 25 to 50 breaths per minute; for children between 1 and 5 years old, between 20 and 40 breaths per minute. Any respiratory rate outside those limits was considered abnormal; 3) HR: for infants between 0 and 1 year old, the normal heart rates were between 100 and 160 beats per minute (bpm); for children between 1 and 3 years old, between 90 and 150 bpm; and for children between 3 and 5 years old, between 80 and 140 bpm. Any heart rate outside those limits was considered abnormal; and 4) Temperature: Any measurement exceeding 38.0 degrees Celsius was considered abnormal.

4. **Sample size calculation**

To calculate the minimum sample size required to detect a difference of 3% in mortality between critically ill children under the IMPALA system compared to usual care, we used the following formula for comparing two proportions:

$$n=\frac{{{(Z}_{\alpha/2}+ Z_{\beta})}^{2} \times(p_{1}\left( 1-p_{1} \right)+ p_{2}\left( 1- p_{2} \right))}{{(p_{1}- p_{2})}^{2}}$$

Where:

- $n$ is the sample size per group.
- $Z_{\alpha/2}$ is the Z-score for the desired significance level (i.e., for 95% confidence, $Z_{\alpha/2}=1.96$).
- $Z_{\beta}$ is the Z-score for the desired power (e.g., for 80% power, $Z_{\beta}=0.8$).
- $p_{1}$ is the mortality rate for the IMPALA monitoring system group.
- $p_{2}$ is the mortality rate for the usual care group.
- $(p_{1}- p_{2})$ is the difference in mortality rates (3% or 0.03).

**R code:**

**# Parameters**

alpha <- 0.05 # Significance level (for 95% confidence)

beta <- 0.2	# Power (for 80% power)

p1 <- 0.1	# Mortality rate for the monitoring system group (e.g., 10%)

p2 <- 0.07	# Mortality rate for the usual care group (e.g., 7%)

delta <- 0.03 # Difference in mortality rates (3%)

**# Z-scores**

Z_alpha <- qnorm(1 - alpha / 2) # Z-score for alpha/2 (two-tailed)

Z_beta <- qnorm(1 - beta)	# Z-score for beta

**# Sample size calculation**

n <- ((Z_alpha + Z_beta)^2 * (p1 * (1 - p1) + p2 * (1 - p2))) / delta^2

**# Display the result**

n_per_group <- ceiling(n) # Round up to the next whole number

n_per_group

We applied the previously mentioned formula to determine the minimum sample size needed to detect a 10% difference in the occurrence of critical illness events (CIEs).

**# Parameters**

alpha <- 0.05 # Significance level (95% confidence)

beta <- 0.2	# Power (80%)

p1 <- 0.1	# Occurrence rate in one group (e.g., 10%)

p2 <- 0.2	# Occurrence rate in the other group (e.g., 20%)

delta <- abs(p1 - p2) # Difference in proportions (10%)

**# Z-scores**

Z_alpha <- qnorm(1 - alpha / 2) # Z-score for two-tailed test

Z_beta <- qnorm(1 - beta)	# Z-score for power

**# Sample size calculation**

n <- ((Z_alpha + Z_beta)^2 * (p1 * (1 - p1) + p2 * (1 - p2))) / delta^2

**# Round up to the next whole number**

n_per_group <- ceiling(n)

**# Display the result**

n_per_group

5. **Critical Illness Event (CIE) definition**

A CIE was defined as the occurrence of any life-threatening event or life-saving intervention during a hospital stay (yes/no). CIE types (i.e., Respiratory, Circulatory, Neurological, Infectious, or Other) and their respective life-threatening conditions were defined as shown in **Supplemental Table 2**.

**Supplemental Table 2. Critical Illness Event type and life-threatening condition definitions**

| **Type of CIE** | **Life-threatening conditions** |
| --- | --- |
| **Respiratory** | Start or increase of respiratory support: oxygen or CPAP |
|  | (Non)Invasive ventilation: bag & mask ventilation or intubation |
|  | Start or increase bronchodilator support |
| **Circulatory** | Transfusion of blood (products) |
|  | Intravenous fluid bolus of 10ml/kg or more |
|  | Start or increase continuous/intermittent inotropic support (IV/IM adrenalin) |
|  | Cardio-pulmonary resuscitation (CPR): resuscitation setting involving chest compressions |
| **Neurological** | Decrease in Blantyre Coma Score of 1 point or more |
|  | Convulsion requiring anticonvulsants |
| **Infectious** | Sepsis: clinical suspicion of sepsis that has led to the collection of a new blood culture and/or start or change in antibiotic treatment |
|  | Start of anti-malarial treatment |
| **Other** | Objectified hypoglycaemia requiring correction (IV or enteral) |
|  | Unplanned admission to the (P)ICU |
|  | Unplanned surgical procedure (including chest drains) |

6. **Disability-adjusted life years (DALYs) calculation**

DALYs were calculated for each child as the sum of years of life lost due to premature mortality (YLL) and years of life lived with disability (YLD) as shown by Formula 1.

$DALY=YLL+YLD$	Formula (1)

The YLL was calculated by subtracting the age of death ($a$) from the life expectancy at birth ($L$) in Malawi (i.e., 62.9 years). Children who did not die, had zero YLL.

$YLL=L-a$	Formula (2)

The YLD was calculated by multiplying the sum of disability weights ($DW$) related to the CIEs during HDU stay by the duration of the HDU stay in days converted to years ($L_{2}$). Disability weights related to CIEs were based on the Global Burden of Disease report as shown in **Supplemental Table 3**^1^.

$YLD=DW \times L_{2}$	Formula (3)

**Supplemental Table 3. Critical Illness Event type and disability weights**

| **Type of CIE** | **Disability weights** | **Definition** |
| --- | --- | --- |
| **Respiratory** | 0.133 | Disability weight for uncontrolled asthma |
| **Circulatory** | 0.224 | Disability weight for cardiac dysrhythmias |
| **Neurological** | 0.552 | Disability weight for severe epilepsy |
| **Infectious** | 0.133 | Disability weight for infectious disease: acute episode, severe |
| **Other** | 0.128 | Disability weight for severe wasting |

Global Burden of Disease Collaborative Network. Global Burden of Disease Study 2019 (GBD 2019) Disability Weights. 2020. DOI:10.6069/1W19-VX76.

**Supplemental Table 4. Distribution of post-IMPALA diagnostic categories used as proxies for CIE types to derive weighted disability weights for population-level DALYs averted extrapolation**

| **Location** | **Type of CIE** | **Count** | **Proportion** |
| --- | --- | --- | --- |
| ZHC paediatric ward | Respiratory | 1664 | 0.27 |
| ZHC paediatric ward | Circulatory | 91 | 0.01 |
| ZHC paediatric ward | Neurological | 221 | 0.04 |
| ZHC paediatric ward | Infectious | 1454 | 0.23 |
| ZHC paediatric ward | Other | 2825 | 0.45 |
| ZCH-HDU | Respiratory | 351 | 0.48 |
| ZCH-HDU | Circulatory | 26 | 0.04 |
| ZCH-HDU | Neurological | 119 | 0.16 |
| ZCH-HDU | Infectious | 118 | 0.16 |
| ZCH-HDU | Other | 122 | 0.17 |
| SLH paediatric ward | Respiratory | 265 | 0.24 |
| SLH paediatric ward | Circulatory | 0 | 0.00 |
| SLH paediatric ward | Neurological | 9 | 0.01 |
| SLH paediatric ward | Infectious | 414 | 0.37 |
| SLH paediatric ward | Other | 438 | 0.39 |

**Additional details on the application and interpretation of extrapolated DALYs averted at the population level**

### **6.1 TMLE-based approach**

The total number of DALYs averted was estimated by applying the average treatment effect (ATE) of the IMPALA intervention on DALYs per child (compared with standard care) to the total number of children admitted during the intervention period.

**Total DALYs averted = DALYs ATE per child × number of children admitted (post-IMPALA)**

Using this approach:

- ZCH-HDU: 5.4 × 736 = 3,974 DALYs averted
- SLH paediatric ward: 1.0 × 1,126 = 1,126 DALYs averted

This approach directly estimates the causal effect of IMPALA on DALYs as a single outcome, capturing both mortality and morbidity within one unified counterfactual framework. These estimates represent total population-level health gains over the intervention period.

### **6.2 Decomposition approach**

DALYs averted can also be estimated by decomposing outcomes into mortality (YLL) and morbidity (YLD) components using counterfactual (TMLE-based) ATE estimates applied to intermediate endpoints.

**Total DALYs averted = (number of children admitted (post-IMPALA) × mortality reduction (percentage points) × remaining life expectancy) + (number of children admitted (post-IMPALA) × CIE reduction (percentage points) × weighted disability weight)**

**Mortality component (YLL)**

Assuming a remaining life expectancy of 60 years:

- ZCH paediatric ward:

YLL = 6,255 × 0.019 × 60 = 7,130

- ZCH-HDU:

YLL = 736 × 0.098 × 60 = 4,328

- SLH paediatric ward:

YLL = 1,126 × 0.016 × 60 = 1,081

**Morbidity component (YLD from CIEs)**

CIE-specific ATEs and disability weights were used:

- CIE ATE (paediatric wards): 0.255
- CIE ATE (ZCH-HDU): 0.471

Thus:

- ZCH paediatric ward:
  YLD = 6,255 × 0.255 × 0.147* = 235
- ZCH-HDU:
  YLD = 736 × 0.471 × 0.203* = 70
- SLH:
  YLD = 1,126 × 0.255 × 0.134* = 39

* Weighted disability weights were derived using the distribution of post-IMPALA diagnostic categories by location as shown in (**Supplemental Table 4**) mapped to Global Burden of Disease disability weights available in **Supplemental Table 3**.

**Total DALYs averted = YLL + YLD**

Using this approach:

- ZCH paediatric ward: 7,130 + 235 = 7,365 DALYs averted
- ZCH-HDU: 4,328 + 70 = 4,398 DALYs averted
- SLH: 1,081 + 39 = 1,120 DALYs averted

Note that there is overlap between the ZCH paediatric ward and ZCH-HDU populations. Accordingly, DALYs averted in the paediatric ward alone represent a lower bound of total DALYs averted at ZCH, whereas the combined ward and HDU estimate represents an upper bound.

**Interpretation and comparison**

Both approaches rely on counterfactual (causal) effect estimation and reflect lifetime health losses averted per event, but are scaled to observed post-IMPALA admissions to obtain population-level estimates. However, they differ in structure:

- DALYs averted could not be estimated for the ZCH pediatric ward using the TMLE-based approach because CIEs were not measured in this location. In contrast, the decomposition approach permits extrapolation of CIE impact estimates from the SLH pediatric ward, allowing DALYs averted to be approximated for the ZCH pediatric ward.
- The TMLE-based approach directly estimates DALYs as a single composite outcome under intervention versus standard care. This approach accounts for the joint dependence of mortality and morbidity in the population.
- The decomposition approach estimates causal effects on mortality and CIEs separately, which are then translated into DALY components. While this approach has the advantage that it shows the separate contributions of mortality and morbidity to DALYs, it makes additional assumptions (e.g., fixed life expectancy, additivity, and independence between components). As a result, it relies more strongly on structural assumptions about how mortality and CIE changes translate into DALYs and may not fully reflect their joint causal pathway.

7. **IMPALA system costs**
   7.1 **Zomba Central Hospital**

Costs calculations for paediatric ward at Zomba Central Hospital for 9 devices and 1 server-unit.

1. Initial Costs:
   - Monitors: $1,500 × 9 = $13,500
   - Server Unit: $3,000 × 1 = $3,000
   - Installation Fee: $1,500
   - Total Initial Costs: $13,500 + $3,000 + $1,500 = $18,000
2. Annual Service Costs:
   - Monitors: $365 × 9 = $3,285
   - Server Unit: $365 × 1 = $365
   - Total Annual Service Costs: $3,285 + $365 = $3,650
3. Service Costs Over 7 Years:
   - $3,650 × 7 = $25,550
4. Total Costs Over Lifetime:
   - Initial Costs + Service Costs: $18,000 + $25,550 = $43,550
5. Costs Per Year:
   - $43,550 ÷ 7 = $6,221.43
6. Costs Per Month:
   - $6,221.43 ÷ 12 = $518.45

Summary

- Total Cost Over 7 Years for the whole installation: $43,550
- Cost Per Year for the whole installation: $6,221.43
- Cost Per Month for the whole installation: $518.45
- Average children admitted to the paediatric ward yearly: 5,000
- Average costs of the IMPALA system per child (2024): $6,221.43/ 5,000 = **$1.24**

### 7.2 **St. Luke’s Hospital**

Costs calculations for paediatric ward at St. Luke’s Hospital for 6 devices and 1 server-unit.

1. Initial Costs:
   - Monitors: $1,500 × 6 = $9,000
   - Server Unit: $3,000 × 1 = $3,000
   - Installation Fee: $1,500
   - Total Initial Costs: $9,000 + $3,000 + $1,500 = $13,500
2. Annual Service Costs:
   - Monitors: $365 × 6 = $2,190
   - Server Unit: $365 × 1 = $365
   - Total Annual Service Costs: $2,190 + $365 = $2,555
3. Service Costs Over 7 Years:
   - $2,555 × 7 = $17,885
4. Total Costs Over Lifetime:
   - Initial Costs + Service Costs: $13,500 + $17,885 = $31,385
5. Costs Per Year:
   - $31,385 ÷ 7 = $4,483.57
6. Costs Per Month:
   - $4,483.57 ÷ 12 = $373.63

Summary

- Total Cost Over 7 Years for the whole installation: $31,385
- Cost Per Year for the whole installation: $4,483.57
- Cost Per Month for the whole installation: $373.63
- Average children admitted to the paediatric ward yearly: 1,000
- Average costs of the IMPALA system per child (2024): $4,483.57/ 1,000 = **$4.48**
- **Average** **costs of the IMPALA system per child (2024) = ($4.48 + $1.24)/ 2 = $2.86**

8. **Direct medical costs (one-day inpatient costs)**
   8.1 **Zomba Central Hospital**

The costs of one day at Zomba Central Hospital were estimated based on the annual HDU expenditure at the Zomba Central Hospital in 2023 (**Supplemental Table 4**) divided by the number of HDU beds (i.e., 16), and subsequently divided by 365 days (i.e., bed-day-based approach). Costs were adjusted to 2024 Malawi inflation using the Consumer Price Index (CPI) (<https://www.nsomalawi.mw/publications/economy/consumer-price-indices-september-2024>) (i.e., September 2024 = 196.6, February 2023 = 134.7)

- **Bed-day-based cost estimation (2023):** ($128,662/16)/ 365 = $22
- **Bed-day-based cost estimation (2024):** $22 x (196.6/ 134.7) = $32

The bed-day-based approach assumes: 1) the care provided per bed-day is consistent across different patients; 2) any fluctuations in staffing levels or resource usage are averaged out over time; and 3) the average occupancy rate is a reasonable proxy for full utilization.

The admission-based approach estimated the one-day HDU costs by dividing the annual HDU expenditure at the Zomba Central Hospital in 2023 by the total number of admissions in 2023 (i.e., 4,340). The admission-based approach thus assumes an HDU duration of 1 day, which may be not the case for critically ill children.

- **Admission-based cost estimation (2023):** $128,662/4,340 = $30
- **Admission-based cost estimation (2024):** $30 x (196.6/ 134.7) = $44

**Supplemental Table 5. Cost items used to estimate a one-day HDU stay based on the annual HDU expenditures at the Zomba Central Hospital (2023)**

| **Items** | **Expenditure (in $)** | **Percentage (%)** |
| --- | --- | --- |
| Personnel | 65,477.15 | 50.9 |
| Utilities | 11,175.51 | 8.7 |
| Food | 3,821.69 | 3.0 |
| Medication | 31,617.65 | 24.6 |
| Vaccine accessories | 3,676.47 | 2.9 |
| Maintenance-Medical equipment | 661.76 | 0.5 |
| Maintenance-building | 4,199.10 | 3.3 |
| Referrals | 2,518.38 | 2.0 |
| Rentals | 5,514.71 | 4.3 |
| **Annual HDU expenditure** | **128,662.42** | **100%** |

A total of 4,340 children were admitted to the HDU Zomba Central Hospital in 2023. HDU includes 8 adult beds. Two children (related or unrelated) could be admitted to a single HDU adult bed, meaning that 16 paediatric beds are available.

8.2 **St. Luke’s Hospital**

The cost of one day at St. Luke's Hospital was calculated by dividing each patient's total hospital bill (in MWK) by their length of stay, converting the result to $ using the most recent available 2024 PPP exchange rate of 1 $ = 382 MWK (https://data.worldbank.org/), and then averaging the per-day cost across all patients.

- **Patient-level cost estimation (2024):** $32

9. **Zomba Central Hospital Caregivers survey**

**Supplemental Table 6. Direct non-medical cost and indirect cost information reported by caregivers**

| **Cost information, MWK** | **N= 150** | |
| --- | --- | --- |
|  | **Mean (SD), MWK** | **Median (IQR), MWK** |
| **Direct non-medical cost information** |  |  |
| Days spent with a child at HDU | 4 (5) | 2 (3) |
| Travel days to or from the hospital | 9 (7) | 8 (8) |
| One-trip costs to or from the hospital | 5,127 (3379) | 4,000 (4000) |
| Transportation costs per one-day HDU stay (MWK) | 22,154 (25877) | 12,250 (26000) |
| Amount spent on food during HDU stay (MWK) | 3,512 (5069), n=57 | 2,000 (4000), n=57 |
| Food costs per on-day HDU stay (MWK) | 1,181 (1050), n=57 | 1,000 (2000), n=57 |
| Amount spent on accommodation during HDU stay (MWK) | 17 (132), n=57 | 0 (0), n=57 |
| Accommodation costs per one-day HDU stay (MWK) | 4 (33), n=57 | 0 (0), n=57 |
| Any other incurred costs during the HDU stay | 2,351 (3187), n=57 | 1,000 (4000), n=57 |
| Any other incurred costs per one-day HDU stay | 1,018 (1529), n=57 | 375 (1625), n=57 |
| **Indirect cost information** |  |  |
| Amount of income loss due to missed work during HDU stay | 373 (1637), n=51 | 0 (0), n=51 |
| Absenteeism costs per one-day HDU stay | 294 (1501), n=51 | 0 (0), n=51 |

Note: Data based on a survey conducted with 150 caregivers of critically ill children admitted to ZCH-HDU between January and May 2023. MWK = Malawian Kwacha. 1$ = 382 MWK (2024) based on the Purchasing Power Parity conversion factor from the World Bank. N=total number of interviewed caregivers; n=complete responses. HDU = High-dependency unit.

10. **Overview: healthcare utilization items and unit prices**

The table below shows an overview of healthcare utilization and unit prices used in the cost-effectiveness analysis.

**Supplemental Table 7. Healthcare utilization items and unit prices**

| **Resource item** | **Unit prices**  **$, 2024** | **Unit prices MWK, 2024** | **Source** |
| --- | --- | --- | --- |
| IMPALA monitoring system (cost per child) | 3 | 1,146 | GOAL 3 |
| One-day HDU stay (bed-based) | 32 | 12,224 | Zomba Central Hospital |
| One-day paediatric ward stay | 32 | 12,224 | St. Luke’s Hospital |
| One-day HDU stay transportation | 58 | 22154 | Caregivers survey (n=150) |
| One-day HDU stay food | 3 | 1181 | Caregivers survey (n=150) |
| One-day HDU stay accommodation | 0.01 | 4 | Caregivers survey (n=150) |
| One-day HDU stay any other costs | 3 | 1018 | Caregivers survey (n=150) |
| One-day HDU stay absenteeism from work | 1 | 294 | Caregivers survey (n=150) |

Note: Non-medical costs based on a survey conducted with 150 caregivers of critically ill children admitted to ZCH-HDU between January and May 2023. MWK = Malawian Kwacha. 1$ = 382 MWK (2024) based on the Purchasing Power Parity conversion factor from the World Bank. HDU = High-dependency unit.

11. **Targeted maximum likelihood estimation (TMLE)**

Targeted maximum likelihood estimation (TMLE) was used to estimate the Average Treatment Effect (ATE) between exposed and unexposed groups. The ATE represents the average effect of the IMPALA monitoring system on the entire population of children, regardless of their actual use of the device.^1,2^

TMLE estimates the conditional mean of the outcome (Y) based on intervention (A) and potential confounders (X) while accounting for the probability of A given the observed X^1,2^. This method served as the main analysis and followed three main steps:^3^

1. Fitting g-computation models for mortality, CIEs, and DALYs and costs using relevant covariates to estimate ATEs.
2. Fitting a propensity score model to match pre- and post-IMPALA cohorts and calculate inverse probability weights.
3. Updating the ATE estimates based on propensity scores.

TMLE provides unbiased estimates even if the G-computation (i.e., the outcome model) or the propensity score model is misspecified, assuming all relevant confounders are captured (i.e., no unmeasured confounding).^1,2^

G-computation models were fitted, including mortality, CIE, and DALYs as dependent variables, and exposure to IMPALA plus a set of covariates that were significantly different at admission, along with those deemed relevant by researchers. For Zomba Central Hospital's paediatric ward, ATEs were adjusted for age, sex, HIV status, multimorbidity, malaria, and the interaction between cohort groups and malaria. For Zomba Central Hospital's HDU, ATEs were adjusted for age, sex, abnormal respiratory rate, abnormal temperature, Blantyre Coma Scale, multimorbidity, monitoring by IMPALA devices, malaria, and the interaction between cohort groups and malaria. The interaction term was included because malaria cases were more frequent in the post-IMPALA period, potentially confounding the association between cohort groups and the outcomes, mortality, and CIE. For St. Luke’s Hospital's paediatric ward, adjustments were made for age, sex, HIV status, and multimorbidity.

The propensity score models were fitted with cohort groups as the dependent variable and a set of covariates that provided the best balance between cohorts. The balance between matched cohorts was assessed using density plots, which compared the distribution of propensity scores. A greater overlap in the distributions indicated more comparable groups.^4^ For the Zomba Central’s paediatric ward data, the propensity score model included age, sex, HIV status, multimorbidity, and month of admission. For the Zomba Central’s HDU data, good propensity score distribution overlap was obtained by fitting the propensity score model with age, sex, HIV status, vital signs, Blantyre coma scale, and month of admission as covariates. For the St. Luke’s paediatric ward, the propensity score model included age, sex, month of admission, and multimorbidity.

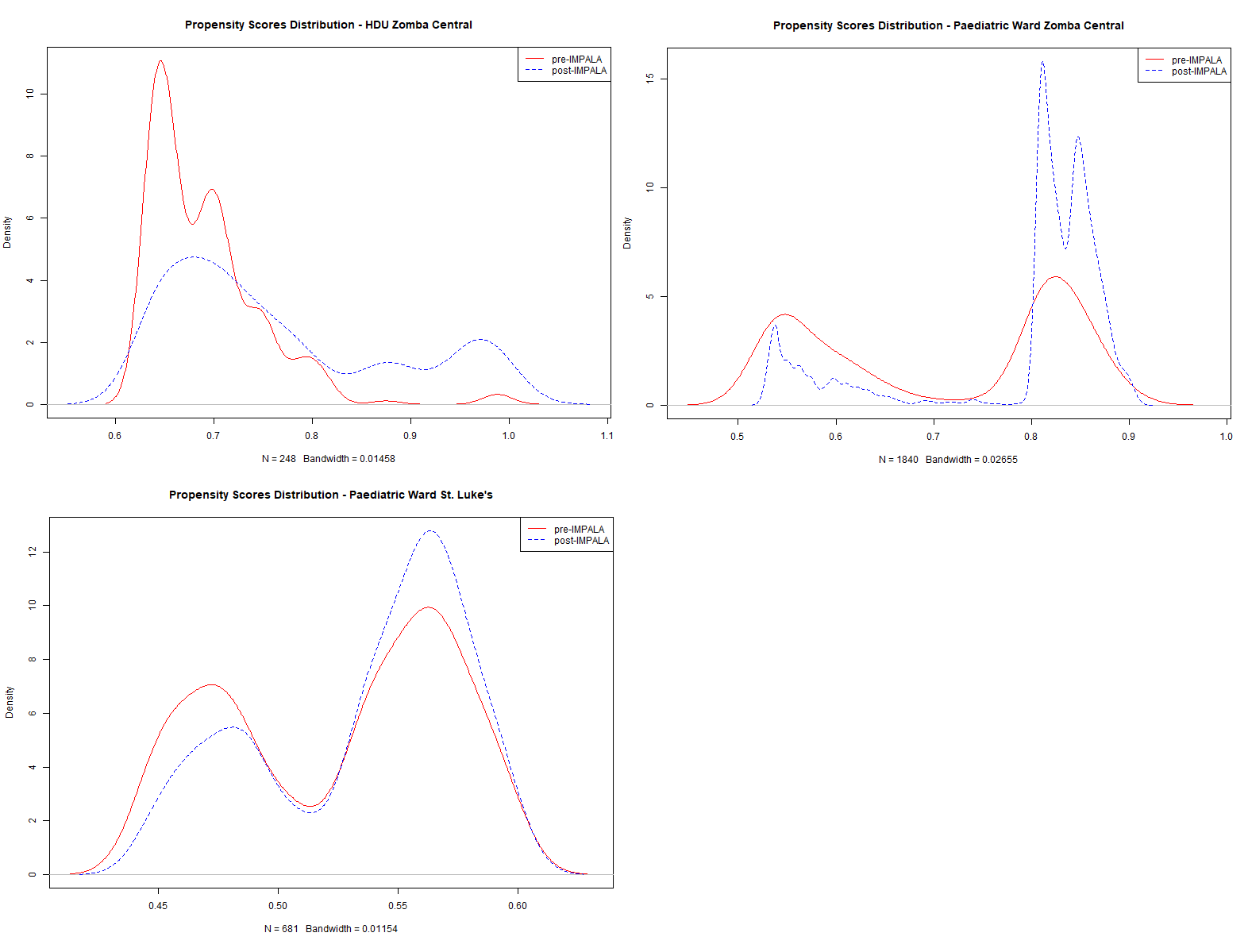

**Supplemental Figure 1. Density plots.** Density plots showing relatively good propensity score distributions overlap between pre- and post-IMPALA cohorts in the three study settings. A good overlap means that pre-IMPALA and post-IMPALA data were comparable for the analysis. HDU: High-Dependency Unit.

1 Kreif N, Grieve R, Radice R, Sekhon JS. Regression-adjusted matching and double-robust methods for estimating average treatment effects in health economic evaluation. Health Serv Outcomes Res Method. 2013;13(2):174-202. doi:10.1007/s10742-013-0109-2

^2^ van der Laan MJ. Targeted Maximum Likelihood Based Causal Inference: Part I. *Int J Biostat*. 2010;6(2):2. doi:10.2202/1557-4679.1211

^3^ Luque-Fernandez MA, Schomaker M, Rachet B, Schnitzer ME. Targeted maximum likelihood estimation for a binary treatment: A tutorial. *Statistics in Medicine*. 2018;37(16):2530-2546. doi:10.1002/sim.7628

^4^ Markoulidakis A, Taiyari K, Holmans P, et al. A tutorial comparing different covariate balancing methods with an application evaluating the causal effects of substance use treatment programs for adolescents. *Health Serv Outcomes Res Method*. 2023;23(2):115-148. doi:10.1007/s10742-022-00280-0

12. **Missing data descriptives: Zomba Central Hospital – High-Dependency Unit data**

**Supplemental Table 8. Missing data**

|  | **Pre-IMPALA, n=248** | | **Post-IMPALA, n=736** | |
| --- | --- | --- | --- | --- |
| **Variable** | **Count** | **Percentage** | **Count** | **Percentage** |
| Age | 0 | 0 | 0 | 0 |
| Sex | 0 | 0 | 0 | 0 |
| Weight | 28 | 11.3% | 178 | 24.2% |
| Abnormal oxygen saturation | 0 | 0 | 0 | 0 |
| Abnormal respiratory rate | 0 | 0 | 0 | 0 |
| Abnormal heart rate | 0 | 0 | 0 | 0 |
| Abnormal temperature | 0 | 0 | 0 | 0 |
| HIV reactive or exposure | 0 | 0 | 0 | 0 |
| Blantyre coma score, mean (SD) | 0 | 0 | 0 | 0 |
| Hematologic/Oncologic | 0 | 0 | 0 | 0 |
| Respiratory | 0 | 0 | 0 | 0 |
| Gastrointestinal diseases | 0 | 0 | 0 | 0 |
| Neurological | 0 | 0 | 0 | 0 |
| Renal/Cardiovascular | 0 | 0 | 0 | 0 |
| Other | 0 | 0 | 0 | 0 |
| Systemic or Severe Infection/Inflammation | 0 | 0 | 0 | 0 |
| Malaria | 0 | 0 | 0 | 0 |
| Malnutrition | 0 | 0 | 0 | 0 |
| Length of stay | 0 | 0 | 2 | 0.3% |
| Mortality | 0 | 0 | 0 | 0 |
| CIEs | 0 | 0 | 0 | 0 |
| DALY | 0 | 0 | 2 | 0.3% |
| Healthcare costs | 0 | 0 | 2 | 0.3% |
| Societal costs | 0 | 0 | 2 | 0.3% |

13. **Missing data descriptives: St. Luke’s Hospital – Paediatric ward data**

**Supplemental Table 9. Missing data**

|  | **Pre-IMPALA, n=930** | | **Post-IMPALA, n=1 126** | |
| --- | --- | --- | --- | --- |
| **Variable** | **Count** | **Percentage** | **Count** | **Percentage** |
| Age | 0 | 0 | 0 | 0 |
| Sex | 0 | 0 | 0 | 0 |
| HIV reactive or exposure | 1 | 0.1% | 1 | 0.1% |
| Hematologic/Oncologic | 0 | 0 | 0 | 0 |
| Respiratory | 0 | 0 | 0 | 0 |
| Gastrointestinal diseases | 0 | 0 | 0 | 0 |
| Neurological | 0 | 0 | 0 | 0 |
| Other | 0 | 0 | 0 | 0 |
| Systemic or Severe Infection/Inflammation | 0 | 0 | 0 | 0 |
| Malaria | 0 | 0 | 0 | 0 |
| Malnutrition | 0 | 0 | 0 | 0 |
| Length of stay | 0 | 0 | 0 | 0 |
| Mortality | 0 | 0 | 0 | 0 |
| CIEs | 249 | 26.8% | 354 | 31.4% |
| DALY | 249 | 26.8% | 354 | 31.4% |
| Healthcare costs | 0 | 0 | 0 | 0 |
| Societal costs | 0 | 0 | 0 | 0 |

14. **Supplemental Figures**

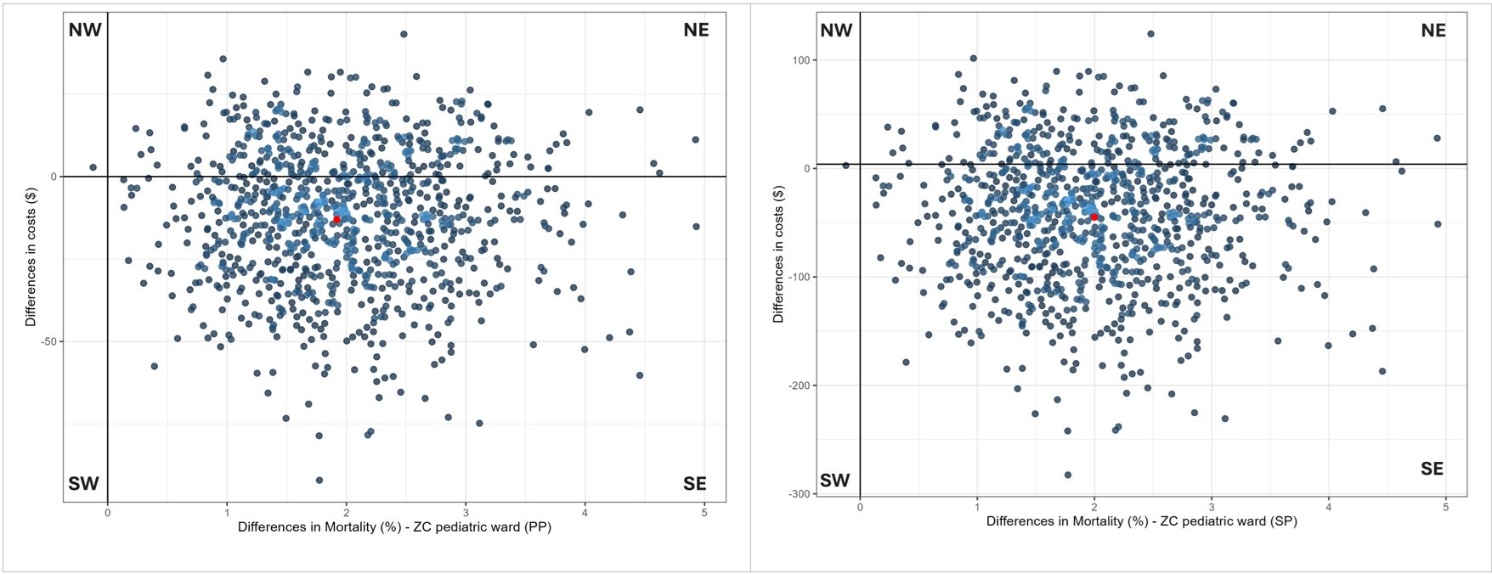

**Supplemental Figure 1. Cost-effectiveness (CE) planes for mortality from a provider (PP) and societal perspective (SP) at the paediatric ward of Zomba Central Hospital.** The CE-planes display the incremental cost-effectiveness ratio (ICER, red dot) and the distribution of 1,000 bootstrapped cost-effect pairs (blue dots).

NE = Northeast (IMPALA monitoring system is more costly and more effective than standard care). The change in mortality was multiplied by -1 as a positive value indicates improvement in outcomes.

SE = Southeast (IMPALA monitoring system is less costly and more effective than standard care).

SW = Southwest (IMPALA monitoring system is less costly and less effective than standard care).

NW = Northwest (IMPALA monitoring system is more costly and less effective than standard care).

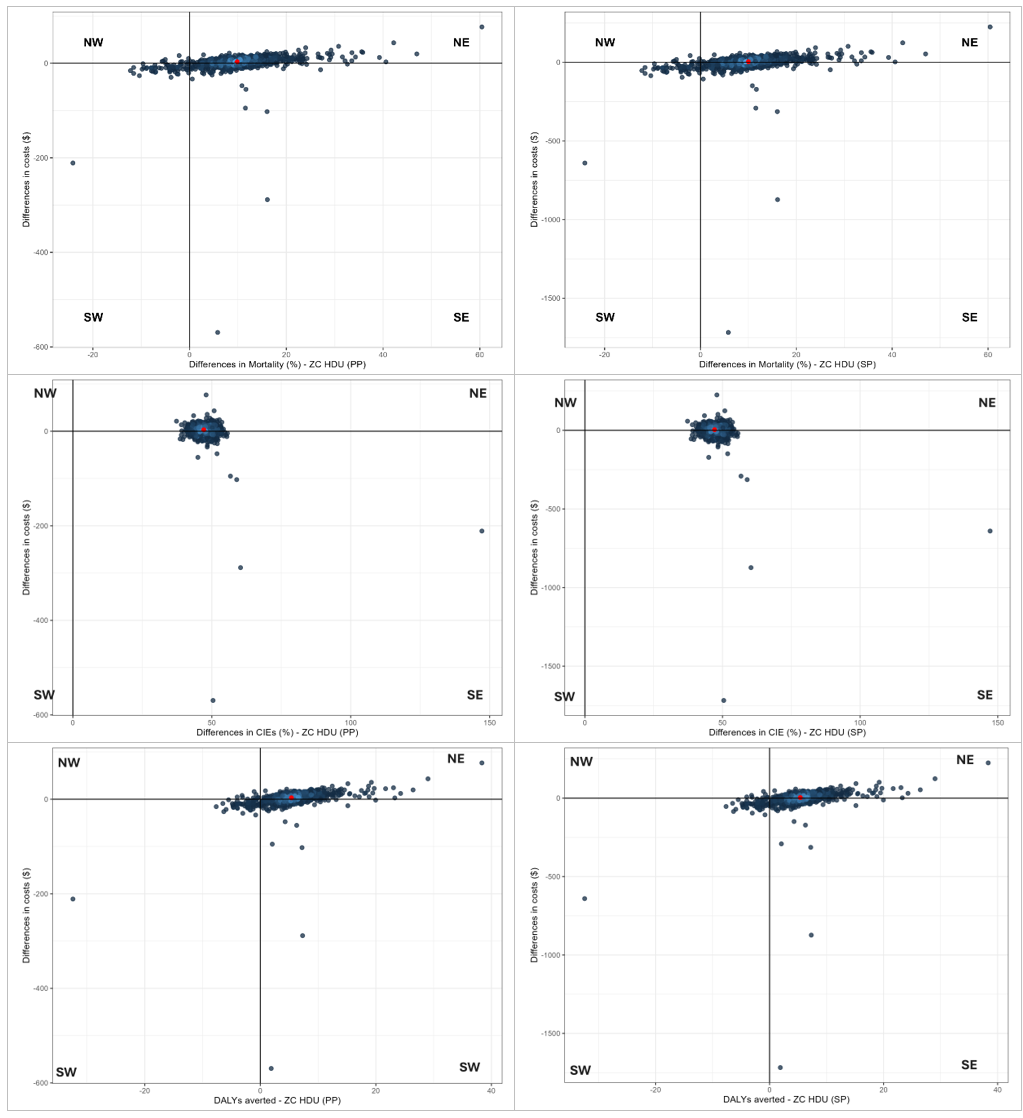

**Supplemental Figure 2. Cost-effectiveness (CE) planes for different outcomes from a provider (PP) and societal perspective (SP) at the High-Dependency Unit of Zomba Central Hospital.** The CE-planes display the incremental cost-effectiveness ratio (ICER, red dot) and the distribution of 1 000 bootstrapped cost-effect pairs (blue dots). The left-hand panels show the results from a PP perspective, the right-hand panels show the SP perspective; the top row shows the mortality CE-planes, middle row critical illness events (CIEs), bottom row. Disability-adjusted life-year (DALY). The changes in mortality, CIEs, and DALYs were multiplied by -1 as positive values indicate improvement in outcomes.

NE = Northeast (IMPALA monitoring system is more costly and more effective than standard care).

SE = Southeast (IMPALA monitoring system is less costly and more effective than standard care).

SW = Southwest (IMPALA monitoring system is less costly and less effective than standard care).

NW = Northwest (IMPALA monitoring system is more costly and less effective than standard care).

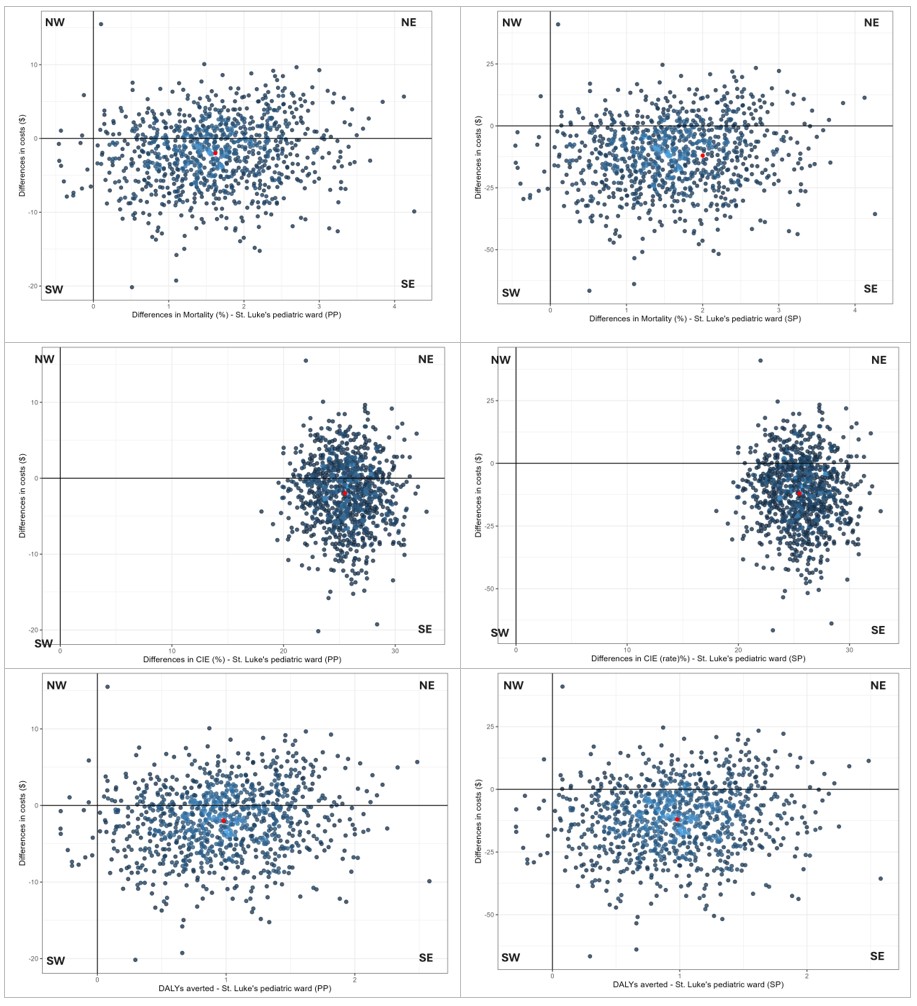

**Supplemental Figure 3. Cost-effectiveness (CE) planes for different outcomes from a provider (PP) and societal perspective (SP) at the paediatric ward St. Luke’s Hospital.**

The left-hand panels show the results from a PP perspective, the right-hand panels show the SP perspective; the top row shows the mortality CE-planes, middle row critical illness event (CIE), bottom row Disability-adjusted life-year (DALY). The CE-planes display the incremental cost-effectiveness ratio (ICER, red dot) and the distribution of 1 000 bootstrapped cost-effect pairs (blue dots). The changes in mortality, CIEs, and DALYs were multiplied by -1 as positive values indicate improvement in outcomes.

NE = Northeast (IMPALA monitoring system is more costly and more effective than standard care).

SE = Southeast (IMPALA monitoring system is less costly and more effective than standard care).

SW = Southwest (IMPALA monitoring system is less costly and less effective than standard care).

NW = Northwest (IMPALA monitoring system is more costly and less effective than standard care).

15. **Sensitivity analysis**

**Supplemental Table 10. Sensitivity analysis by location**

| **Effect outcome** | **ΔE (95% CI)** | **ΔC, $ (95% CI)** | **ICER** | **Cost-effectiveness plane** | | | | **Probability of cost-effectiveness** | | |
| --- | --- | --- | --- | --- | --- | --- | --- | --- | --- | --- |
|  |  |  |  | **NE** | **SE** | **SW** | **NW** | **WTP**  **$0** | **WTP**  **$224** | **WTP**  **$448** |
| **Paediatric ward, Zomba Central Hospital, n=1 703 per cohort** | | | | | | | | | | |
| Mortality %, PP* | 2.1 (0.8; 3.6) | -22.2 (-53.5; 0.4) | -10.5 | 3 | 97 | 0 | 0 | 1.0 | 1.0 | 1.0 |
| Mortality %, SP* | 2.1 (0.7; 3.6) | -72.6 (-168.4; -5.0) | -34.8 | 1 | 99 | 0 | 0 | 1.0 | 1.0 | 1.0 |
| **High-Dependency Unit, Zomba Central Hospital, n=120 per cohort** | | | | | | | | | | |
| Mortality %, PP* | 1.7 (-4.3; 8.4) | 4.9 (-25.0; 35.5) | 2.8 | 14 | 13 | 26 | 48 | 0.1 | 1.0 | 1.0 |
| Mortality %, SP* | 1.7 (-4.3; 8.4) | 9.0 (-80.9; 101.0) | 5.2 | 45 | 29 | 14 | 13 | 0.1 | 1.0 | 1.0 |
| CIEs %, PP* | 67.4 (56.3; 78.3) | 4.9 (-25.0; 35.5) | 0.1 | 61 | 39 | 0 | 0 | 0.1 | 1.0 | 1.0 |
| CIEs %, SP* | 67.4 (56.3; 78.3) | 9.0 (-80.9; 101.0) | 0.1 | 57 | 43 | 0 | 0 | 0.1 | 1.0 | 1.0 |
| DALYs, PP* | 1.1 (-2.7; 5.2) | 4.9 (-25.0; 35.5) | 4.6 | 48 | 26 | 13 | 13 | 0.1 | 0.8 | 0.8 |
| DALYs, SP* | 1.1 (-2.7; 5.2) | 9.0 (-80.9; 101.0) | 8.4 | 45 | 29 | 14 | 13 | 0.1 | 0.8 | 0.8 |
| **Paediatric ward, St. Luke’s Hospital, n=745 for mortality, pre-IMPALA n=557 and post-IMPALA n=527 for CIEs and DALYs** | | | | | | | | | | |
| Mortality %, PP* | 1.8 (0.2; 3.5) | -0.8 (-10.1; 8.2) | -0.4 | 44 | 54 | 1 | 0 | 0.6 | 1.0 | 1.0 |
| Mortality %, SP* | 1.8 (0.2; 3.5) | -8.3 (-36.2; 19.1) | -4.3 | 28 | 71 | 1 | 0 | 1.0 | 1.0 | 1.0 |
| CIEs %, PP* | 27.1 (22.2; 32.4) | -3.3 (-13.9; 8.0) | -0.1 | 28 | 72 | 0 | 0 | 0.9 | 1.0 | 1.0 |
| CIEs %, SP* | 27.1 (22.2; 32.4) | -15.9 (-47.8; 18.5) | -0.6 | 18 | 82 | 0 | 0 | 1.0 | 1.0 | 1.0 |
| DALYs, PP* | 1.1 (0.12; 2.12) | -3.3 (-13.9; 8.0) | -2.9 | 28 | 71 | 1 | 0 | 0.9 | 0.9 | 0.9 |
| DALYs, SP* | 1.1 (0.12; 2.12) | -15.9 (-47.8; 18.5) | -14.2 | 18 | 80 | 1 | 0 | 1.0 | 1.0 | 0.9 |

CIEs = critical illness events during hospital stay (yes/no). DALYs = disability-adjusted life-years.

*Effect outcomes were multiplied by -1 to ensure an interpretable cost-effectiveness plane, as lower mortality indicates a better effect, ensuring a more interpretable cost-effectiveness plane.

PP = provider perspective

SP = societal perspective

ΔE = Average Treatment Effect on the Treated (ATET) for effect outcomes.

ΔC = Average Treatment Effect on the Treated for costs. CI = confidence interval.

ICER = Incremental Cost-Effectiveness Ratio ($ per unit of effect gained).

NE = Northeast (IMPALA is more costly and more effective than standard care).

SE = Southeast (IMPALA is less costly and more effective than standard care) -> cost-saving

SW = Southwest (IMPALA is less costly and less effective than standard care).

NW = Northwest (IMPALA is more costly and less effective than standard care).

WTP = Willingness-to-pay thresholds based on 0.5 and 1 Gross Domestic Product per capita in Malawi 2024.
